# Integrated Host–Microbe Signatures Differentiate Respiratory Infection from Incidental Pathogen Carriage

**DOI:** 10.1101/2025.10.27.25338902

**Authors:** Emily C. Lydon, Padmini Deosthale, Abigail Glascock, Hoang Van Phan, Christina M. Osborne, Matthew K. Leroue, Jawara Allen, Eran Mick, Brandie D. Wagner, Joseph L. DeRisi, Lilliam Ambroggio, Peter M. Mourani, Charles R. Langelier

## Abstract

Accurately distinguishing lower respiratory tract infection (LRTI) from incidental pathogen carriage (IPC) is clinically challenging. The host immunologic and microbial factors that define the states of LRTI and IPC are poorly understood. We performed host-microbe metatranscriptomic profiling of tracheal aspirate from 326 mechanically ventilated children with clinically adjudicated LRTI (n=207), IPC (n=70), or non-infectious acute respiratory illnesses (n=49). In the airway microbiome, LRTI was characterized by reduced alpha diversity and taxonomic richness, while IPC was characterized greater total bacterial abundance, enrichment in respiratory anaerobes and increased metabolic activity. In terms of host response, patients with LRTI exhibited a distinct lower airway transcriptional signature of innate and adaptive immune activation compared to those with IPC, who had similar transcriptional profiles as uninfected controls. Mediation analyses suggested that the airway microbiome influences the host response to pathogens. An integrated host-microbe metatranscriptomic classifier discriminated LRTI from IPC and controls with an AUC=0.89 (95% confidence interval (CI) 0.85–0.92). The single gene *FABP4,* when combined with alpha diversity, performed similarly, and FABP4 protein alone achieved an AUC=0.88 (95% CI 0.82–0.93). Together, our findings reveal distinct ecological and immunologic archetypes that define LRTI and IPC, and support data-driven, biology-informed LRTI diagnostics that incorporate host and microbe.

## INTRODUCTION

The upper and lower airways harbor robust microbial communities which together represent the human respiratory microbiome^1^. While primarily comprised of commensal bacteria during states of health, it has been well established that respiratory pathobionts, or microbes with the potential to cause disease, can be incidentally carried within the airway microbiome without eliciting signs or symptoms of infection^2^. This phenomenon of ‘colonization’ or incidental pathogen carriage (IPC), which involves a dynamic relationship between pathobiont, microbiome and host immune response, remains incompletely understood.

IPC frequently complicates the management of acute respiratory illness by confounding accurate lower respiratory tract infection (LRTI) diagnosis. For instance, in patients hospitalized for respiratory failure, the underlying cause often remains unclear for days, as both infectious and non-infectious conditions can present with overlapping clinical features. LRTI diagnostic tests that rely almost exclusively on pathogen detection cannot differentiate between true infection and IPC, nor discern the presence of a non-infectious etiology^3,4^. As a result, detection of any potential pathogen, particularly in the setting of respiratory failure, can lead to an LRTI diagnosis, even in the absence of true infection^5,6^. This contributes to unnecessary antimicrobial use and missed opportunities to diagnose and treat alternative causes of respiratory failure, such as cardiac conditions or autoinflammatory diseases^7–9^.

While IPC occurs across the age spectrum, the incidence is highest in children compared to adults^10–12^. For instance, an estimated 33-90% of children incidentally carry *Streptococcus pneumoniae* in the airway, compared to <5% of adults^13–16^. Similarly, *Moraxella catarrhalis* colonizes the nasopharynx in 30-100% of infants but only 1-5% of adults^17–19^. Viral IPC is also far more common among children; based on population surveillance studies, an estimated 25% of asymptomatic young children incidentally carry at least one viral pathogen, in contrast to only 2% of adults^20–22^.

The high baseline rates of respiratory IPC in children may be further increased in the setting of hospitalization and critical illness. The physiological disturbances of critical illness, including disruption of epithelial barriers, immune dysregulation, and introduction of endotracheal tubes, can reshape the respiratory microenvironment, promoting shifts in microbial community composition and facilitating colonization by opportunistic pathogens^23,24^. Among patients who require mechanical ventilation for non-infectious indications, airway colonization with potentially pathogenic organisms occurs in nearly half within the first 24 hours^25,26^.

Despite its clinical relevance and frequent occurrence, the host and microbial factors that distinguish IPC from LRTI remain incompletely understood. No studies have yet evaluated IPC in the context of the lower airway microbiome and host response, and tests capable of accurately distinguishing LRTI from IPC do not yet exist. To address these gaps, we studied a prospective cohort of critically ill children hospitalized for acute respiratory failure and performed metatranscriptomic RNA sequencing on lower respiratory samples to simultaneously profile both host and microbe. We identify striking host and microbial biosignatures that distinguish the two states, and then leverage findings to build accurate diagnostic classifiers with the potential to advance acute respiratory illness management.

## RESULTS

### Patient cohort, clinical adjudication, and pathogen detection

Critically ill children with all-cause respiratory failure requiring mechanical ventilation (n=457) were prospectively enrolled at eight U.S. hospitals between 2/2015 and 12/2017 (**Figure S1**).^27–29^ Tracheal aspirate was obtained within 24 hours of intubation and stored in an RNA stabilizing agent. High-quality RNA sequencing data capturing both the respiratory microbiome and host transcriptome were generated on 343 patients.

LRTI cases were identified by structured, retrospective clinical adjudication following ICU discharge performed by ≥2 physicians trained in critical care medicine or infectious diseases using the CDC/NHSN PNU1 criteria^30^. Adjudicators, who had access to all clinical data in the electronic health record and were blinded to metatranscriptomic results^31^, identified 224 patients (65.3%) with LRTI. Alternative, non-infectious etiologies of respiratory failure were adjudicated in 119 (34.7%) patients, and included neurologic conditions, anatomic airway abnormalities, toxin exposures, cardiac conditions, trauma and autoimmune disease. To comprehensively identify respiratory pathogens in the lower airway, a combination of standard-of-care clinical microbiologic testing and respiratory metatranscriptomics was performed. Following this process, 207 patients received a clinical diagnosis of LRTI and had a respiratory pathogen detected (“LRTI” group). Of those with a clear non-infectious cause of respiratory failure, 70 had a pathogen detected (“IPC” group) and 49 patients did not (“CTRL” group). Seventeen patients with clinically adjudicated LRTI but negative microbiologic testing were excluded from the analysis.

We found no differences in sex, race, ethnicity, or comorbidities between groups. Patients in the LRTI group had a younger median age of 0.6 years (interquartile range (IQR) 0.2-2.1), compared to 1.6 years (IQR 0.9-6.4) in the IPC group and 9.5 years (IQR 1.3-14.4) in the CTRL group, reflecting the typical epidemiologic differences in pediatric respiratory failure.^4,32^ Ventilator days and ICU length of stay were slightly longer in LRTI versus IPC, though hospital length of stay was similar and mortality was lower. Antibiotic usage prior to intubation was similar across all groups, and notably, most patients received antibiotics during their hospital course (LRTI 99.0%, IPC 91.4%, CTRL 83.7%).

Respiratory syncytial virus (RSV) was the most common respiratory pathogen in the cohort, identified in 52.7% of the LRTI group and 12.9% of the IPC group (P<0.001) (**Figure 1**). Other pathogens that statistically differed in prevalence between groups included human metapneumovirus (LRTI 6.8%, IPC 0.0%, P=0.02), *Haemophilus influenzae* (LRTI 30.0%, IPC 17.1%, P=0.04), and *Pseudomonas aeruginosa* (LRTI 1.9%, IPC 10.0%, P=0.01). Several other pathogens were identified at similar rates between groups, including rhinovirus, *Moraxella catarrhalis*, *Staphylococcus aureus*, and *Streptococcus pneumoniae*. Co-detection of both bacteria and viruses was common, comprising 59.4% of LRTI cases and 25.7% of IPC cases.

**Figure 1.**
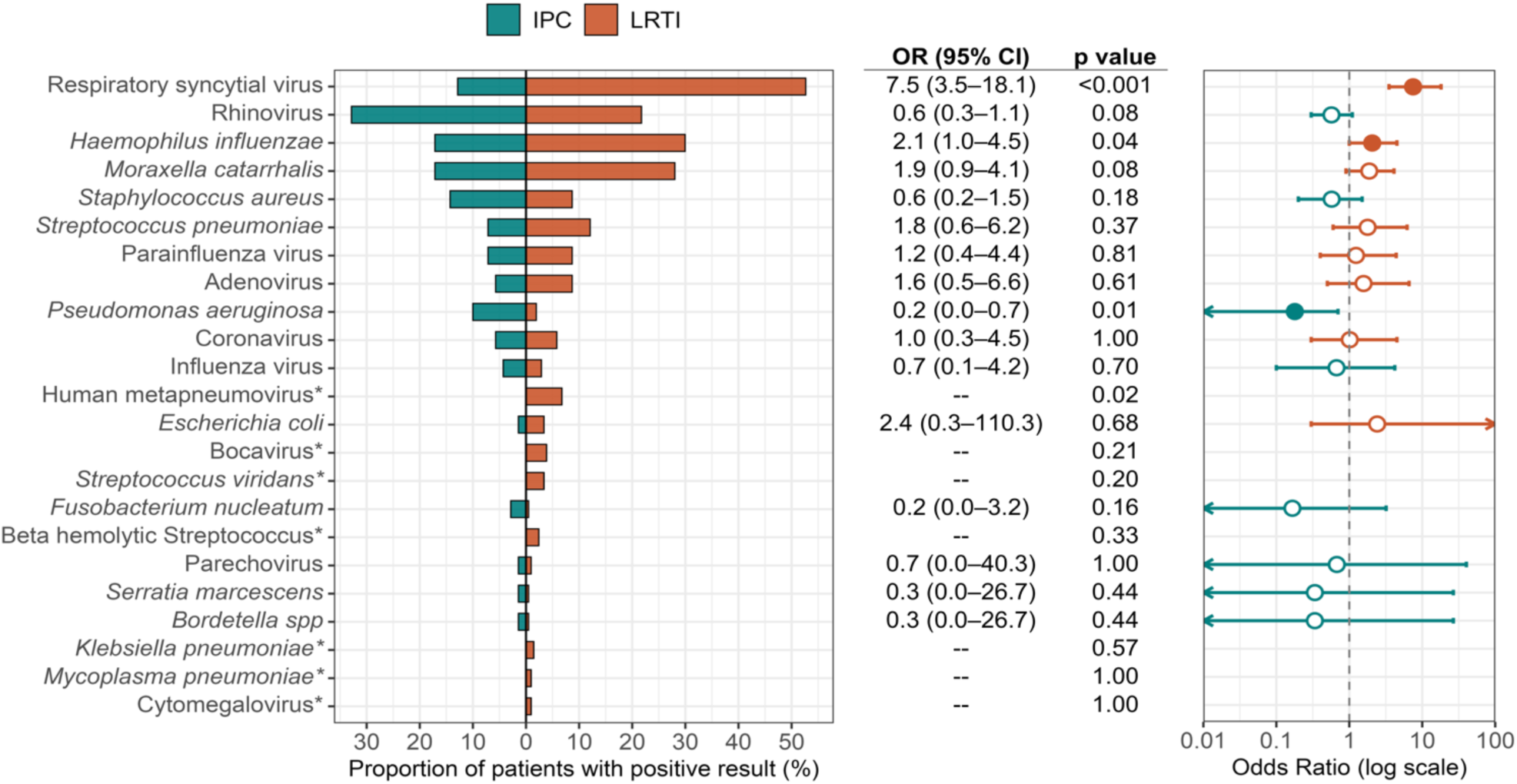
Distribution of pathogens among patients with clinically adjudicated lower respiratory tract infection (LRTI) or incidental pathogen carriage (IPC). Bar plot demonstrating the proportion of participants in the LRTI (n = 207, orange) and IPC (n = 70, teal) groups with each detected pathogen as determined by combined clinical testing and metatranscriptomics. Odds ratio (OR) with 95% confidence interval (CI) tabulated and plotted on the right. Filled circles represent statistically significant pathogens based on P<0.05. Arrows indicate where the lower or upper confidence interval exceeds the plotting area. Pathogens detected only once in the entire cohort were excluded from plotting. Statistical significance for between-group differences was assessed with Fisher’s exact test.

### The respiratory microbiome differs between LRTI, IPC, and controls

We first sought to compare the composition and function of the lung microbiome between LRTI and IPC, hypothesizing that both biologically relevant and diagnostically useful distinctions may exist. Alpha diversity demonstrated notable differences, with LRTI characterized by a lower Shannon Diversity Index (SDI) compared to either IPC (P_adj_=2.8e-8) or CTRL (P_adj_=2.6e-14). In contrast, SDI did not differ between IPC and CTRL groups (P_adj_=0.59) (**Figure 2a**). Similarly, we found that community richness (total number of unique species in the lung microbiome) was significantly higher in both IPC and CTRL groups compared to those with LRTI (P_adj_=1.2e-11 and 1.3e-9, respectively) (**Figure 2b**). Community composition also differed between LRTI, IPC, and CTRL groups based on the Bray-Curtis index (P<0.001) (**Figure 2c**). Despite these distinct microbiome archetypes, the detected pathogen was typically the most abundant microbe in the airway microbiome in both LRTI and IPC (**Figure S2**).

**Figure 2.**
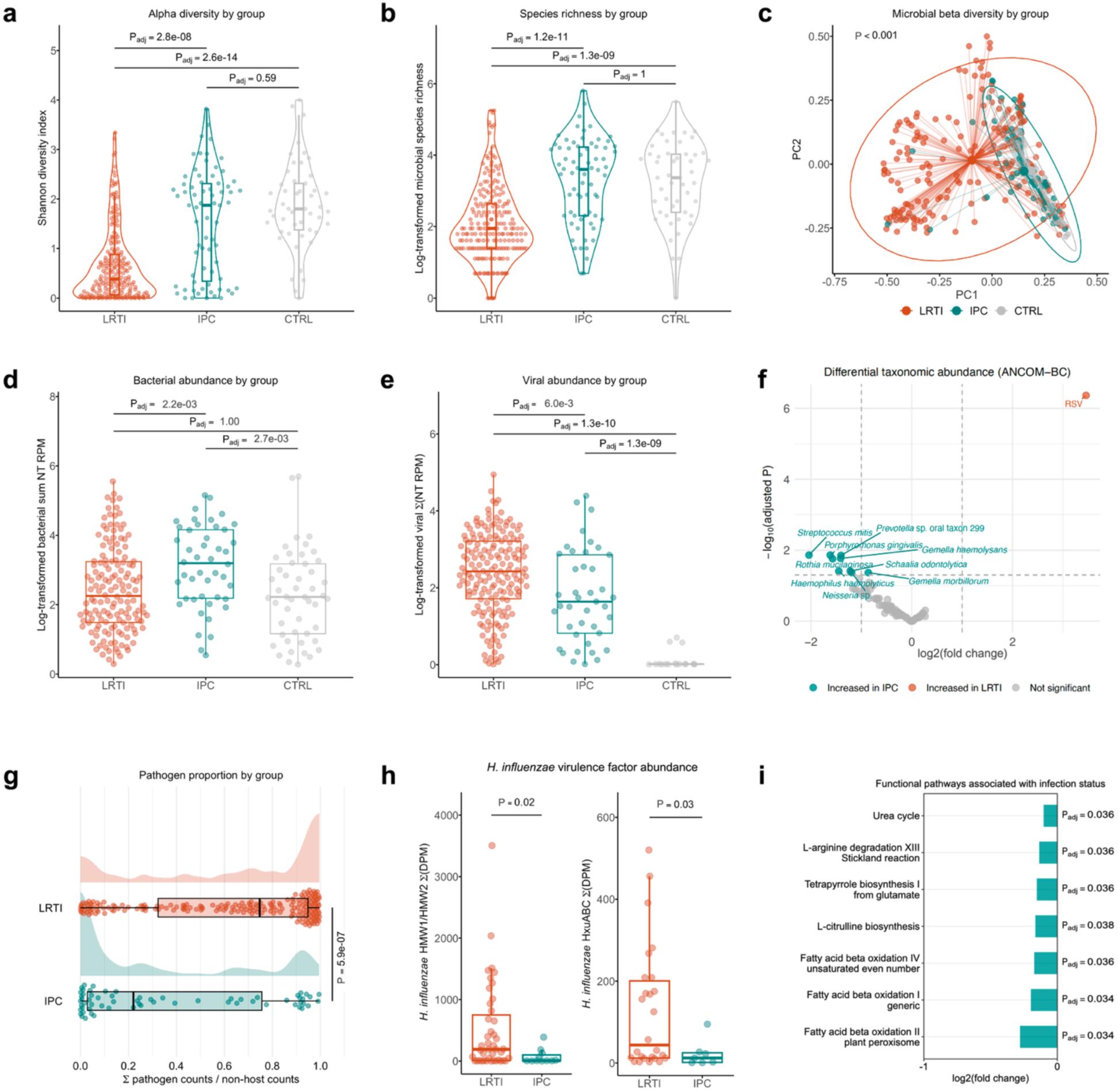
The lower airway microbiome differs between infection states. **a.** Shannon diversity index (SDI) of the lung microbiome in patients with LRTI (n=207, orange), IPC (n=70, teal), or CTRL (n=49, gray). **b.** Species richness of the lung microbiome by group. **c.** Principal-coordinate analysis of Bray–Curtis distances demonstrating differences in community composition between groups; p value calculated by PERMANOVA. **d.** Lower airway total bacterial abundance measured in reads per million (RPM). **e.** Lower airway total viral abundance measured in RPM. **f.** Differentially abundant taxa in the lower airway microbiome identified by ANCOM-BC. **g.** Proportion of pathogen-assigned reads represented in the lower airway microbiome. **h.** Expression of *Haemophilus influenzae* virulence factors *HMW1/2* and *HxuABC* across groups. **i.** Log₂-fold change in HUMAnN3-derived bacterial metabolic pathways found to be significantly different (P_adj_<0.05) between groups.

We next compared total bacterial abundance (measured in reads per million, RPM) between groups. Surprisingly, total bacterial abundance was highest in IPC compared to both LRTI (P_adj_=2.2e-3) and CTRL (P_adj_=2.7e-3) (**Figure 2d**). In contrast, total viral RPM was highest in LRTI, while the IPC group exhibited an intermediate state with greater viral abundance compared to the CTRL group (P_adj_=1.3e-9) (**Figure 2e**). Sensitivity analyses subsetting by viral-bacterial co-detection did not markedly change the observed relationships between LRTI, IPC, and CTRL groups (**Figure S3**).

We next examined pathogen abundance and found that rhinovirus RPM was significantly higher in LRTI compared to IPC (P=0.047), while mean RSV abundance trended higher in LRTI, but the difference did not reach statistical significance (P=0.060) (**Figure S4**). *H. influenzae* abundance did not differ between groups (P=0.84), while *M. catarrhalis* RPM was unexpectedly higher in IPC compared to LRTI (P=9.1e-3).

To further characterize the microbiome features of LRTI and IPC, we carried out differential taxonomic abundance analysis using ANCOM-BC^33^. While RSV was the only microbe with significantly greater abundance in LRTI, we found that IPC was enriched with classically commensal taxa including *Prevotella*, *Neisseria*, *Porphyromonas*, and *Streptococcus* species (**Figure 2f, Supp. Data 1**). We subsequently evaluated pathogen burden, quantified as the proportion of implicated pathogen reads in the microbiome, and found that LRTI was characterized by a significantly greater pathogen burden compared to IPC (63.4% versus 36.6% mean pathogen proportion, P=5.9e-7) (**Figure 2g)**.

We hypothesized that virulence factor expression might differ between LRTI and IPC, and thus carried out an exploratory assessment using the MetaVF database^34^. We found that the expression of two *H. influenzae* virulence factors, *HMW1/2* and *HxuABC,* were higher in LRTI (P=0.02 and P=0.03, respectively) (**Figure 2h, Figure S5, Supp. Data 2**). *HMW1/2* are adhesin proteins that facilitate *H. influenzae* adherence to the respiratory epithelium^35^, and *hxuABC* is a specialized ATP-binding cassette transporter for iron acquisition from the host^36^.

We additionally assessed microbiome functional differences by profiling metabolic pathways using HUMAnN^37^. We found that compared to LRTI, IPC was characterized by higher expression of metabolic pathways essential for fatty acid beta-oxidation, citrulline biosynthesis, and arginine degradation (**Figure 2i, Supp. Data 3**). Taken together, our findings suggested that IPC is characterized by a more diverse, taxonomically rich, abundant, and metabolically active respiratory microbiome compared to LRTI.

### Host airway transcriptional responses distinguish LRTI from IPC and controls

We next tested the hypothesis that the pulmonary host response would differ between LRTI, IPC, and CTRL groups by evaluating the lower airway transcriptome. Principal component analysis demonstrated that LRTI was characterized by a distinct transcriptional signature compared to IPC or CTRL groups (PERMANOVA P_adj_=0.002), which did not differ (P_adj_=0.25) (**Figure 3a, Figure S6**). This finding was underscored by hierarchical clustering of the top 20 most differentially expressed (DE) genes between LRTI and CTRL groups, which generally separated LRTI from non-LRTI cases, though several IPC cases clustered among LRTI cases (**Figure 3b**). IPC and CTRL patients did not clearly separate based on hierarchical clustering.

**Figure 3.**
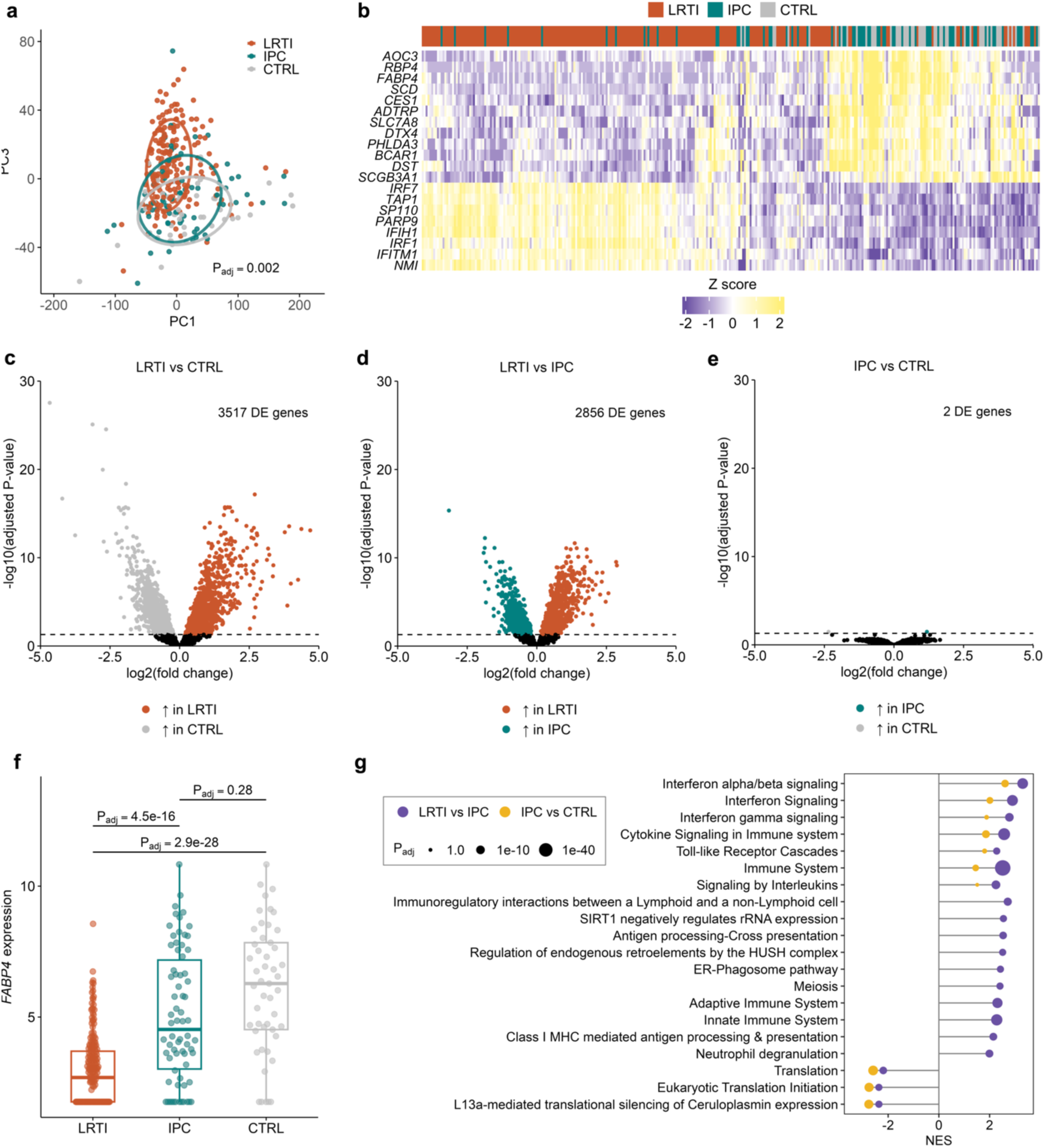
Lower airway host transcriptional responses distinguish LRTI from IPC and controls. **a.** Principal component analysis (PCA) of the lower respiratory tract transcriptome. Adjusted P value calculated by PERMANOVA between LRTI and non-LRTI groups. **b.** Heat map demonstrating hierarchical clustering of patients in each group (LRTI, IPC, CTRL) based on the top 20 differentially expressed (DE) genes between LRTI and CTRL groups. Color bar indicates normalized, Z-score scaled expression of each gene. **c.** Volcano plot of DE genes between LRTI and CTRL, with Benjamini-Hochberg (BH) adjusted P values <0.05 colored. **d.** Volcano plot highlighting DE genes between LRTI and IPC. **e.** Volcano plot of DE results comparing IPC versus CTRL. **f.** Normalized *FABP4* expression across groups, with Benjamini-Hochberg adjusted P values from the DE analyses. **g.** Gene-set enrichment analysis (GSEA) highlighting immune pathway enrichment in IPC compared to LRTI (purple) and IPC compared to CTRL (yellow). Top 20 DE pathways shown for the LRTI/IPC comparison, then overlaid with the IPC/CTRL comparison, showing only significant pathways (P_adj_<0.05). Point size scales inversely with P_adj_ value.

To better understand host gene expression differences between the three groups at a more granular level, we performed pairwise differential expression analyses, adjusting for age and sex. We identified distinct host signatures that differentiated LRTI from IPC and CTRL groups, with 3517 and 2856 DE genes, respectively (**Figure 3c**, **Figure 3d, Supp. Data 4**). In contrast, IPC and CTRL groups demonstrated minimal differences, with only 2 DE genes (**Figure 3e, Supp. Data 4**). Among the genes DE between LRTI and both CTRL and IPC groups, we noted that *FABP4*, which is expressed in macrophages and encodes a lipid chaperone that modulates leukotriene stability^38^, was a clear outlier in both fold change and statistical significance (**Figure 3f).**

To characterize biological pathways encompassing the DE genes, we carried out gene set enrichment analyses (GSEA) (**Figure 3g, Supp. Data 5)**. Canonical infection-related pathways, including interferon signaling, antigen presentation, adaptive immune signaling, and neutrophil degranulation, were all upregulated in LRTI versus IPC, as expected. However, we also noted that interferon signaling pathways were upregulated in IPC compared to CTRL patients, albeit to a lesser extent,

Given that interferon signaling is a central feature of the anti-viral host immune response, we hypothesized that the immunologic features of IPC may differ between viral and bacterial pathogens. To investigate this, we performed differential expression analyses within patients who had viral (n=223) or bacterial (n=195) pathogens detected. Aligning with our primary composite analysis, both viral and bacterial LRTI were characterized by distinct airway transcriptional signatures with respect to the CTRL group (3424 and 3140 DE genes, respectively) (**Figure 4a, Supp. Data 6**). Compared to IPC, viral and bacterial LRTI also exhibited distinct host signatures, although with fewer DE genes (1860 and 1991, respectively). Few transcriptomic differences were observed between IPC and CTRL groups, although a subtle signature of 29 DE genes, primarily interferon-stimulated genes (ISGs, e.g., *ISG15, IFIH1, OAS3*) characterized viral IPC (**Figure 4b**). In contrast, there were zero DE genes between bacterial IPC and CTRL groups, suggesting fundamental differences in immune activation between patients incidentally carrying viral versus bacterial pathogens.

**Figure 4.**
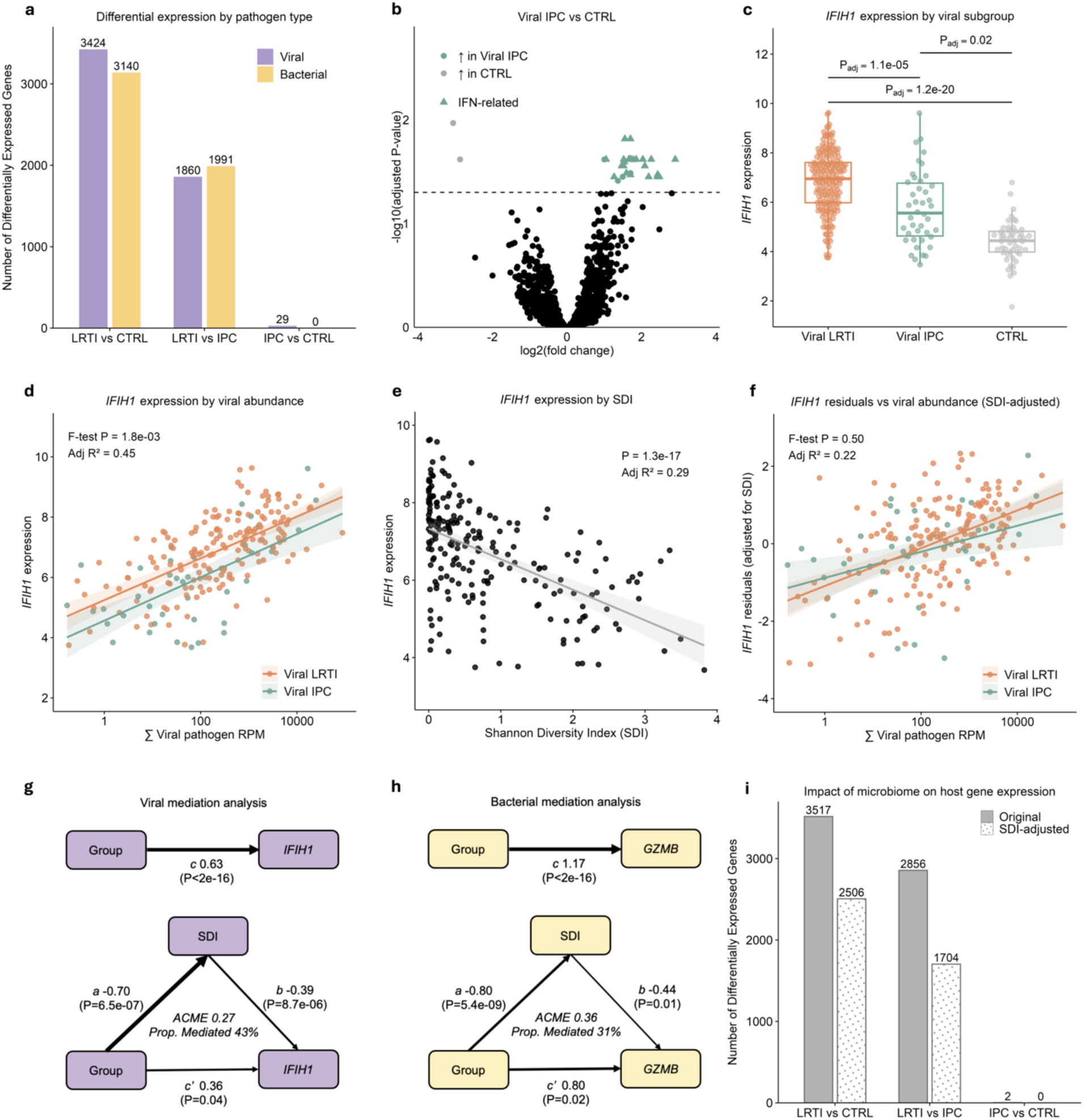
Innate immune activation differs between LRTI and IPC and is modulated by pathogen type, abundance, and microbiome diversity. **a.** Number of differentially expressed (DE) genes between LRTI, IPC, and CTRL stratified by viral (purple) or bacterial (yellow) pathogen class. **b.** Volcano plot highlighting subtle interferon-stimulated gene (ISG) expression signature in viral IPC versus CTRL. **c.** Boxplot of ISG *IFIH1* expression by group. Benjamini-Hochberg (BH) adjusted P values, based on DE analyses, shown. **d.** Linear regression of viral abundance (reads per million, RPM) against *IFIH1* expression for viral LRTI and IPC. Shaded area indicates 95% confidence intervals. **e.** Negative correlation between Shannon diversity index (SDI) and *IFIH1* expression in viral subgroups. **f.** Linear regression between viral abundance and *IFIH1* expression, after adjusting for SDI. **g.** Mediation analysis illustrating proportion of group effect mediated by SDI in viral cases. **h.** Analogous mediation analysis for *GZMB* in bacterial cases. **i.** Number of DE genes (P_adj_<0.05) between pairwise LRTI, IPC and CTRL comparisons, with and without adjustment for SDI.

We next examined expression of individual genes classically associated with anti-viral and anti-bacterial defense. The viral subgroups demonstrated a gradation in ISG expression^39^, ranging from highest in viral LRTI to lowest in CTRL (**Figure 4c)**. Given the similar pattern with viral abundance in our microbiome analysis (**Figure 2e**), we hypothesized that the differences in interferon signaling might simply be related to viral load differences between groups. A regression of ISG expression against viral RPM showed that ISG expression did positively correlate with interferon expression, as expected (adjusted R^2^=0.45), though interestingly, when stratified by group, IPC patients demonstrated a proportionally attenuated response compared to those with LRTI for any given viral load (P=1.8e-3 for the ISG *IFIH1*) (**Figure 4d**). When age was added as a covariate, this finding did not change (P=2.0e-3), and other ISGs (*ISG15*, *IFI44*) exhibited the same pattern (**Figure S7**).

While these findings could be due entirely to intrinsic differences in innate immune response activation between individuals, we considered the possibility that the lung microbiome might modulate, at least to some extent, inflammatory gene expression in the setting of pathogen exposure. To investigate this, we evaluated the relationship between SDI and ISG expression and found that as lung microbiome diversity increased, ISG expression decreased (adjusted R^2^=0.29, regression P=1.3e-17) (**Figure 4e**). Furthermore, we found that after adjusting for SDI, between-group differences in the relationship between viral load and ISG expression disappeared (P=0.50), suggesting that the lung microbiome modulates anti-viral host responses (**Figure 4f**). Mediation analysis suggested that lower SDI was independently associated with higher *IFIH1* expression, and that microbiome composition may partially mediate the relationship between viral pathogen presence and interferon signaling, explaining ∼43% of the group effect on *IFIH1* expression (**Figure 4g**).

We performed a parallel analysis focused on bacterial LRTI and IPC (**Figure S8**). In contrast to our observations with viral pathogens, the expression of canonical anti-bacterial innate immunity genes (*GZMB*^40^, *CD64*^41^, and *TLR1*^42^) remained relatively constant across a range of bacterial pathogen loads (e.g., for *GZMB*, adjusted R^2^=0.07). However, as with viral pathogens, the bacterial IPC group exhibited consistently lower innate immunity gene expression compared to the LRTI group (e.g. for *GZMB*, P_adj_=3.0e-04). Applying the same mediation analysis to *GZMB* demonstrated that lower lung microbiome alpha diversity was independently associated with higher innate immunity gene expression, explaining ∼31% of the group effect on *GZMB* expression (**Figure 4h**).

Lastly, we assessed the impact of adjusting for SDI in our original pairwise differential expression analyses and found that doing so markedly reduced the lower airway transcriptional differences between LRTI and IPC (**Figure 4i, Supp. Data 7**).

### Integration of host and microbial features enables accurate LRTI diagnosis

Having identified such distinct microbiome and host immune response differences between groups, we next sought to translate our findings into proof-of-concept diagnostic tests. Using LASSO regularized regression, we built diagnostic classifiers to distinguish true infection from the alternative clinically encountered states of IPC or non-infectious acute respiratory illness (**Figure 5a**). Given prior work demonstrating the utility of *FABP4* as a pneumonia diagnostic biomarker^43^, we evaluated its performance alone or in combination with alpha diversity. Both *FABP4* and SDI performed well individually, although the combination achieved even better classification performance with an area under the receiver operator curve (AUC) of 0.87 (95% confidence interval (CI) 0.83–0.91) based on 5-fold cross validation (CV). A multi-gene host transcriptional classifier in combination with SDI performed comparably with an AUC of 0.89 (95% CI 0.85–0.92, **Figure 5b, Table S1**) and yielded classifier scores that effectively distinguished LRTI from patients in either the IPC or CTRL groups (**Figure 5c**).

**Figure 5.**
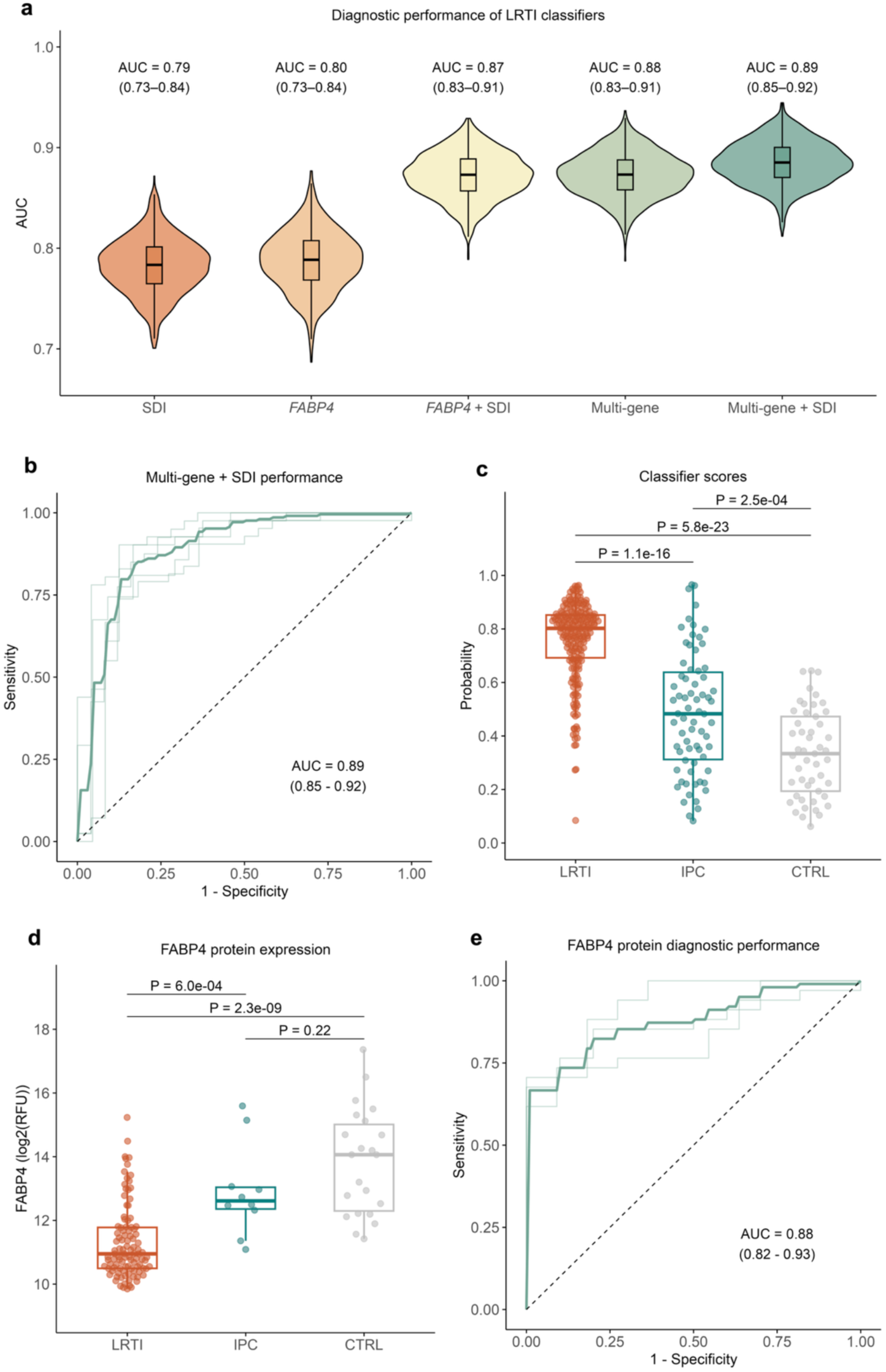
Integration of host and microbial features enable accurate diagnosis of LRTI and differentiation from IPC. **a.** Comparative performance of classifiers for distinguishing LRTI from IPC or CTRL groups. Classifiers based on: Shannon diversity index (SDI), *FABP4, FABP4* + SDI, a multi-gene model derived from LASSO regularized regression of the host transcriptome, or a multi-gene model + SDI. AUC reflects mean across 5-fold cross validation, and 95% CI estimated by bootstrapping. **b.** ROC curve for multi-gene classifier with SDI. **c.** Probability of LRTI based on classifier scores broken down by clinical group. **d.** Lower respiratory FABP4 protein concentration differences between LRTI, IPC and CTRL groups. **e.** Receiver operating characteristic (ROC) curve for FABP4 protein biomarker.

Considering that a single protein biomarker could have distinct practical utility as a clinical diagnostic, we tested whether protein levels of FABP4 could also effectively differentiate LRTI from IPC and CTRL groups in a subset of patients with FABP4 protein measurements from the lower airway (n=134). Indeed, the LRTI group had markedly different levels of FABP4 compared to both IPC (P=6.0e-4) and CTRL (P=2.3e-9) groups (**Figure 5d**). Consistent with our findings at the transcriptional level, no difference in FABP levels between IPC and CTRL groups was observed (**Figure 3f**). Notably, we found that respiratory FABP4 alone performed as well as the integrated host/microbe metatranscriptomic classifier (AUC=0.88, 95% CI 0.82–0.93) (**Figure 5e**), suggesting promise as a clinical biomarker for both LRTI diagnosis and distinguishing LRTI from IPC.

## DISCUSSION

Differentiating LRTI from IPC remains a frequent and unresolved challenge in the care of patients with acute respiratory illness. The resulting diagnostic uncertainty drives antimicrobial overuse and reflects a key gap in our understanding of host-microbe interactions in the lower airway. Here, we deploy metatranscriptomic profiling to holistically characterize the host and microbial features of LRTI and IPC, identifying distinct inflammatory signatures and respiratory microbiome ecology that distinguish these two states of pathobiont existence. Leveraging these findings, we develop host-microbe and practical single biomarker LRTI diagnostic classifiers, offering a path toward more precise, biologically informed diagnostics.

Although the implicated pathogens were frequently the most abundant microbes in the lower airway microbiome in both patients with LRTI and IPC, microbiome alpha and beta diversity, and taxonomic richness were strikingly different. LRTI was marked by a collapse of alpha diversity, reflecting ecologic disruption that is characteristic of infection^27,44,45^, whereas IPC resembled uninfected controls, with a diverse and taxonomically rich community composition. Unexpectedly, total bacterial abundance was greater in IPC than in LRTI or controls, which may be explained by the enrichment of commensal taxa such as *Prevotella*, *Neisseria*, and *Porphyromonas* and reflect a more resilient and balanced barrier microbiota. The IPC state was further characterized by increased expression of diverse metabolic programs (e.g. energy production, fatty-acid β-oxidation, and amino-acid biosynthesis) and higher virulence factor expression. Globally, these findings suggest that IPC is characterized by a more robust, diverse, and metabolically active microbiome, tolerant of carriage but resilient to pathogen invasion.

Host inflammatory gene expression in the lower airway also differed markedly between LRTI and IPC. We found that LRTI elicited a distinct transcriptional signature compared to either IPC or controls, comprised of thousands of DE genes related to innate and adaptive immune signaling. In contrast, the lower airway transcriptome of IPC largely resembled that of controls. Sensitivity analyses demonstrate that while no detectable transcriptional differences existed between bacterial IPC and controls, a subtle signal of interferon-stimulated genes distinguished viral IPC from controls. This pattern is consistent with prior reports of interferon activation during asymptomatic viral carriage^46,47^, but contrasts studies in neonates demonstrating that nasopharyngeal colonization with *M. catarrhalis*, *H. influenzae*, and *S. pneumoniae* correlates with mucosal immune shifts and the future development of asthma^48,49^. The discrepancy may be due to developmental stage (neonates were not included in our study and may be uniquely susceptible as their respiratory microbiomes are being established) or differences in upper versus lower respiratory tract biology^50^.

Regression analyses demonstrated that ISG expression is induced in a viral load-dependent manner, consistent with prior studies^51,52^. Intriguingly, however, ISG activation was consistently diminished in the setting of IPC across a range of viral loads. This suggested that IPC may be characterized by a global attenuation of the pathogen recognition-innate immune activation axis.

Similar regression analyses involving bacterial cases did not demonstrate a dose-dependent relationship with respect to innate immune gene expression, perhaps reflecting fundamental differences in the coupling of bacterial antigens to the transcriptional activation of host innate immune responses. That said, we observed consistently higher expression of anti-bacterial immune genes (e.g., *GZMB*, *CD64*, *TLR1*) in LRTI compared to IPC, across a broad range of pathogen abundance, suggesting a fundamental set point difference between the two states, agnostic to pathogen class.

Our mediation analysis supports a role for the lung microbiome in moderating the intensity of inflammatory responses to potential invading pathogens. These findings, while proof-of-concept in nature, align with murine models in which microbiome disruption amplifies innate inflammatory responses and influenza-associated lung injury^53^. We estimated that microbiome factors explained ∼43% of the relationship between group and inflammatory gene expression in viral cases, and ∼31% in bacterial cases. While an important contribution, this suggests that other mediators (e.g., host genetics, epigenetic modifications, immune memory to related pathogens) primarily account for host responses differences between LRTI and IPC. Regardless, our findings underscore the complex, bi-directional relationship between host and microbe that determines whether a pathobiont will cause invasive disease or co-exist innocuously in a microbial community. We acknowledge that a full mediation analysis and determination of causality and directionality is not possible given the inherent limitations of an observational cohort^54^.

Distinguishing LRTI from IPC and non-infectious acute respiratory illnesses remains a clinical challenge and underscores the need for better diagnostic tests to guide antimicrobial therapy and patient care. We illustrate that simple host and microbial biomarkers can be used independently, or combined, to build clinically translatable diagnostic tests to address this need. For instance, *FABP4* in combination with SDI accurately distinguished children with proven LRTI from those with other causes of acute respiratory failure, including those with IPC, achieving an AUC of 0.87. As sequencing technology becomes more economical and clinically available, the feasibility and cost effectiveness of performing metatranscriptomic analyses will continue to increase^55–57^.

Inflammatory protein biomarkers (e.g., procalcitonin, C-reactive protein) are the most widely clinically available class of host-based infectious disease diagnostics, although they only have modest capability of diagnosing LRTI and have not been shown to effectively discriminate between LRTI and IPC.^58,59^ Thus, we found it promising that FABP4 alone, when measured at the protein level, performed as well as our integrated host/microbe metatranscriptomic model (AUC=0.88), highlighting the potential clinical utility of this single host biomarker for rapid and accurate diagnosis of infection in this cohort.

Our study has several strengths including the incorporation of both host and microbial data using metatranscriptomics, a multicenter design, rigorous and comprehensive adjudication of LRTI and IPC, and a large sample size. Our study also has limitations. We focused exclusively on children because overall they have a higher prevalence of IPC, thus it remains unknown whether our findings are generalizable to adults, or to individuals with less severe respiratory illnesses. The infection and IPC groups differed in age, although we adjusted for this in our analyses. While our study is the largest to date to examine biological differences between infection and IPC, our sample size did limit sub-analyses at the individual pathogen level.

Given the inherent limitations of an observational cohort and cross-sectional study design, our mediation analyses should be considered proof-of-concept and will require validation in a more controlled experimental setting. Longitudinal sampling could help determine whether diversity collapse precedes, accompanies, or follows infection onset, and studies in xenobiotic mice could more effectively establish causal relationships between microbiome and host inflammatory responses in the setting of pathobiont challenge. Finally, future studies are needed to evaluate whether our findings at the host or microbiome level generalize to the upper respiratory tract.

In sum, we find that LRTI and IPC are characterized by distinct biology with respect to both host and microbe, emphasizing that simply detecting a microbe with known pathogenicity in the respiratory tract is insufficient for clinical diagnosis of infection. It is not just the pathogen alone, but its dynamic relationship with the host immune response and airway microbiome, that determines disease. Our study provides fresh insight into the vexing and common challenge of interpreting positive respiratory tests in patients with acute respiratory illnesses and offers a new approach for improving LRTI diagnostic accuracy and limiting antimicrobial overuse.

## METHODS

### Study cohort

We analyzed a prospective multicenter cohort of 457 critically ill children with acute respiratory illnesses requiring mechanical ventilation, who were admitted to eight U.S. intensive care units (ICUs) in the National Institute of Child Health and Human Development’s Collaborative Pediatric Care Research Network (CPCCRN) between February 2015 and December 2017. As described previously in detail,^27–29^ we enrolled children aged 31 days to 18 years who were expected to require mechanical ventilation for at least 72 hours and had tracheal aspirate (TA) sampling performed within 24 hours of intubation. The study was approved by the University of Utah central IRB #00088656.

### Adjudication of infection status and definition of subgroups

Adjudication of LRTI status was carried out retrospectively by study-site clinicians with access to all clinical, laboratory, microbiology, and radiology data available up to the end of admission, without knowledge of metagenomic next-generation sequencing (mNGS) results. Each patient was reviewed independently by two adjudicators with expertise in pediatric infectious disease and/or critical care to determine the presence or absence of clinical LRTI; disagreements were discussed and resolved by a panel. For this study, patients were classified into three groups: 1) LRTI if they were clinically adjudicated as having LRTI and had supportive microbiology, 2) IPC if they were clinically adjudicated as not having LRTI but had positive microbiology, and 3) CTRL if they were clinically adjudicated as not having LRTI and had negative microbiology. Microbiology included standard-of-care clinical microbiology (multiplex polymerase chain reaction (PCR) and semiquantitative bacterial respiratory cultures) and/or metagenomic detection for pathogenic bacteria and viruses (implementing a validated, rules-based computational model, described in detail below)^27,28,44^.

### RNA sequencing

Tracheal aspirate (TA) collected within 24 hours of intubation was mixed equi-volume with DNA/RNA shield (Zymo Research, Cat. No R1100) and stored at -80 °C. Following bead-bashing, RNA or negative control water samples underwent extraction using the Qiagen AllPrep Kit (Qiagen, Cat. No R2145), followed by DNAse treatment. Sequencing libraries were prepared from purified RNA using the NEBNext Ultra II Library Prep Kit (New England Biolabs, Cat. No E7770L) and dual index barcodes. Human ribosomal RNA depletion was carried out prior to library amplification and pooling using the Cas9-based Depletion of Abundant Sequences by Hybridization (DASH) method^60^. Libraries underwent 150-base pair paired-end sequencing on an Illumina NovaSeq 6000 sequencer.

### Measurement of FABP4 protein levels

FABP4 was measured from TA specimens collected within 24 hours of intubation using the SomaScan 7k assay (SomaLogic)^61–63^ in a subset of this cohort. Following collection, TA specimens underwent centrifugation at 4°C at 15,000 × *g* for 5 min, subsequently the supernatant was frozen at −80°C within 30 minutes.

### Taxonomic mapping from RNA-seq data

We employed the CZ ID Illumina mNGS pipeline (v7.1) for taxonomic mapping of microbial sequence data^64,65^. This incorporates initial removal of human reads using Kallisto^66^, adapter sequence trimming with fastp^67^, filtering low quality and low complexity reads using PriceSeq^68^ and the Lempel-Ziv-Welch algorithm, respectively, and a final scrub of any residual human reads using Bowtie2^69^. Taxonomic classification was then performed on both short reads and assembled contigs using the NCBI nucleotide (NT) and nonredundant (NR) databases. Background and batch correction was performed on species level taxon matrices (see below).

### Identification and mitigation of background contaminants

Negative water controls were processed and sequenced alongside the patient samples to enable characterization and subtraction of background contamination. A previously developed negative binomial model^51^ was used to model the distribution of reads of microbial taxa in the negative controls. Mean and dispersion parameters were fit to the data and estimates of the mean were generated for each batch:taxon pair. A single dispersion parameter was generated across all taxa using the MASS package (R, v7.3.58.1). P values were adjusted for multiple testing using the Benjamini-Hochberg False Discovery Rate method. Microbial taxa that were present at a significantly higher average abundance in participant samples than in negative controls (P_adj_<0.05) were retained for downstream analyses. Microbial taxa were included in downstream analysis if they met these criteria: (1) ≥1 hit to the NT database (2) >=1 hit to the NR database (3) a minimum alignment length of 70 bases to the NT database.

### Clinical detection of respiratory pathogens

Standard-of-care clinical respiratory microbiologic testing was performed based on the discretion of the treating clinicians at each study site. Diagnostics included nasopharyngeal swab respiratory pathogen testing by multiplex PCR and/or TA bacterial semiquantitative cultures. Clinical diagnostic tests on samples obtained within 48 hours of intubation were included in the analyses. Microbes reported by the clinical laboratory as representing laboratory, skin, or environmental contaminants, or reported as mixed upper respiratory flora, were excluded.

### Detection of respiratory pathogens by metatranscriptomics

For bacterial taxa that remained after background filtering, we applied an established rules-based model (RBM) to identify potential respiratory pathogens. In two prior studies, the RBM identified 82-96% of clinically-confirmed lower respiratory pathogens compared to standard of care clinical diagnostics, and permitted detection of otherwise missed potential pathogens in >50% of patients with clinically adjudicated LRTI but negative standard testing^27,44^. The RBM operates by first retaining the most abundant species from each mapped genus, and any lower-abundance species within that genus with known pathogenicity in the respiratory tract based on a curated reference list from epidemiologic surveillance studies.^70–73^ Species were then ranked by abundance (reads per million values aligned to NT database, sum NT RPM), limiting to the top 15. The largest drop in abundance among this ranked list was identified, and any species above the largest drop in abundance with known ability to cause LRTI as a potential pathogen were counted as bacterial hits. Viruses detected with an abundance > 0.1 RPM with established human respiratory pathogenicity were subsequently identified.

### Microbial abundance calculations

Microbial abundance/load was approximated for each sample by calculating the sum NT RPM. Statistical comparison of sum NT RPM across groups was performed using the wilcox_test () function (rstatix v0.7.2). Resulting P values were adjusted using the Benjamini-Hochberg False Discovery Rate algorithm via the p_adjust() function of the stats (v4.2.3) package. Generalized linear modeling of these relationships was performed using the glm() function of the stats package, specifying a Gaussian distribution and identity link function, and adjusted for both sex and age. These methods were applied to the entire microbial profile as well as subsets of the profile (e.g. bacterial, viral) based on NCBI lineage data. Differences in abundance (sum NT RPM) of individual species of interest between groups were also performed using this approach.

### Microbiome diversity analyses

Alpha diversity (Shannon Diversity Index, or SDI) was calculated using the diversity() function of the vegan package (v2.6-6.1). Beta diversity (Bray-Curtis dissimilarity) was calculated using the functions vegdist(), betadisper(), permutest() and adonis2() of the vegan package. Principal Coordinate Analysis (PCoA) was performed using the cmdscale() function of the stats package (v4.2.3).

### Differential microbial abundance analysis

Differential abundance analysis was performed using the ANCOM-BC package^33^ (v2.8.1) using a library filter of 0, prevalence filter of 10%, alpha level of 0.05 and a pseudo-count of 1. The analysis was adjusted for age and sex, and P values were adjusted for multiple testing using the Benjamini-Hochberg correction.

### Virulence factor screening

Virulence factors were identified from transcriptomic data using the MetaVF toolkit^34^, its associated virulence factor database VFDB2.0, and BLAST (v2.16.0). Called virulence factors with an associated e-value less than 1e-10 were retained for downstream analysis. Differentially expressed virulence factors were identified using ANCOM-BC^33^ with a library filter of 0, prevalence filter of 5%, alpha level of 0.05 and a pseudo-count of 1.

### Generation of host gene counts

RNA-seq reads were pseudoaligned using Kallisto^66^ against an index consisting of all transcripts associated with human protein coding genes (GRCh38-based). We excluded samples with less than one million exon counts. Gene-level counts were generated using tximport package, with the scaled TPM method^74^. Genes were retained for subsequent analysis if they had at least 10 counts in at least 20% of the samples in the cohort. The gene counts table underwent variance-stabilizing transformation (VST) using the R package DESeq2^75^, and VST-transformed counts were used in the principal component analysis, hierarchical clustering, assessment of individual genes, and classifier development.

### Principal component analysis

Principal component analysis (PCA) was performed on the complete gene expression matrix using the prcomp function. For data visualization, we plotted PC1 versus PC3, which provided the greatest apparent separation in two dimensions, and ellipses depict 68% confidence regions around group centroids. PC1 versus PC2 and PC2 versus PC3 are shown in the supplement. To formally assess group separation, we ran pairwise PERMANOVA (adonis2, vegan package) on Euclidean distances computed from the full expression matrix (i.e., testing differences in group centroids). P values from the pairwise contrasts were adjusted by the Benjamini-Hochberg method, with P_adj_<0.05 considered significant.

### Heat map and hierarchical clustering

For display, we selected the top 20 most significantly differentially expressed genes based in the LRTI and CTRL comparison, based on P_adj_ (see below for DE methods). Each gene was standardized across samples (z-score; mean = 0, SD = 1). Genes were ordered by unsupervised hierarchical clustering using Euclidean distance and complete linkage (ComplexHeatmap defaults). Samples were clustered using correlation distance with Ward.D2 to emphasize similarity of gene expression profiles. Dendrograms were computed for ordering but omitted in final visual panel.

### Differential expression and gene set enrichment analyses

DE analyses were performed with the R package limma-voom on raw gene-level counts^76^. The design matrix included age and sex as covariates, and where noted, Shannon Diversity Index (SDI). Counts were transformed with voom (mean-variance modeling with precision weights) and quantile normalized across samples. Gene-wise statistics used empirical-Bayes moderated two-sided t-tests; multiple testing was accounted for by Benjamini-Hochberg with P_adj_<0.05 considered significant. Where individual genes are displayed (e.g., boxplots for *FABP4*, *IFIH1*, and *GZMB*), the adjusted P values from the respective limma DE analyses are displayed. For pathway analysis, we performed pre-ranked gene set enrichment analysis (GSEA) using ReactomePA (gsePathway) on Reactome gene sets with a minimum pathway size of 10 genes and a maximum size of 1500 genes^77^. All genes from each limma DE comparison, ranked by the limma t-statistic, were included as input. For visualization, we displayed the top 20 DE pathways between LRTI and IPC (all statistically significant with P_adj_<0.05), then overlaid results from the IPC and CTRL comparison, only displaying the significant pathways.

### Regression and mediation analyses

For both viral and bacterial subgroup analyses (run in parallel with identical pipeline), we modeled gene expression using linear regression with log_10_ pathogen abundance (sum NT RPM) and group (LRTI vs IPC) as predictors, including an interaction term (gene expression ∼ log_10_ pathogen abundance * group). Group differences were evaluated using a global F-test comparing this model to the null model (gene expression ∼ log_10_ pathogen abundance), and sensitivity models additionally adjusted for age. For visualization, raw data points are plotted alongside model-predicted regression lines. The same structure was applied for microbiome diversity (gene expression ∼ SDI * group). SDI-adjusted associations were visualized by plotting residuals from the gene expression ∼ SDI model against pathogen abundance with group-specific linear fits. To evaluate modulation of innate host gene expression by diversity, we used the mediation package with SDI as the mediator and included pathogen abundance as a covariate in both models (mediator: SDI ∼ group + log10 pathogen abundance; outcome: expression ∼ group + SDI + log10 pathogen abundance) and calculated average causal mediated effect (ACME) and average direct effect (ADE) with 1000 simulations^78^. The mediation models used simpler additive models to maintain the interpretability of the effect estimates and because the interaction terms were not significant in any of the regression models. Viral outcomes focused on interferon-stimulated genes (*IFIH1*, *ISG15*, *IFI44*) and bacterial outcomes on canonical anti-bacterial defense genes (*GZMB*, *CD64*, *TLR1*).

### Classifier development

Binary classifiers were developed to distinguish LRTI from non-LRTI (IPC + CTRL combined). We performed stratified five-fold cross-validation (same folds re-used across the different models, with a minimum IPC and CTRL counts to keep each fold balanced) and generated out-of-fold predictions for performance assessment. Single-feature models (i.e. *FABP4* gene, FABP4 protein, SDI) used logistic regression, as did FABP4 + SDI. Multi-gene models used LASSO logistic regression on all genes (glmnet), with the regularization parameter lambda selected by internal cross-validation^79^. Non-zero coefficients selected by the LASSO model are provided in Table S1. For performance metrics, the reported AUC reflects the mean AUC of each of the five folds computed with the pROC package^80^, and the confidence intervals were obtained by bootstrapping the out-of-fold predictions with 1000 resamples.

## Data availability

Source data are provided with this paper. The raw fastq files with microbial sequencing reads are available under NCBI BioProject ID: PRJNA748764. Deidentified clinical metadata, host and microbial data, code, and source data files are available at https://github.com/infectiousdisease-langelier-lab/Incidental_pathogen_carriage.

**Table 1:** Demographic and clinical characteristics of the LRTI, IPC, and CTRL groups. Race is indicated as “unknown” if patient declined or was unable to answer, if they selected “other” as an option, or if data were missing. P values compare LRTI and IPC groups. Fisher’s exact test used for categorical variables, and Kruskal-Wallis rank-sum test used for continuous variables. IQR, interquartile range; ICU, intensive care unit. * Indicates statistically significant (P < 0.05)

|  | LRTI (n=207) | IPC (n=70) | P value | CTRL (n=49) |
| --- | --- | --- | --- | --- |
| Female, n (%) | 124 (59.9) | 38 (54.3) | 0.49 | 24 (49.0) |
| Male, n (%) | 83 (40.1) | 32 (45.7) |  | 25 (51.0) |
| Age in years, median (IQR) | 0.6 (0.2-2.1) | 1.6 (0.9-6.4) | <0.001* | 9.5 (1.3-14.4) |
| Race, n (%) |  |  | 0.53 |  |
| White | 122 (58.9) | 39 (55.7) |  | 27 (55.1) |
| Black/African American | 41 (19.8) | 10 (14.3) |  | 11 (22.4) |
| Asian | 9 (4.3) | 4 (5.7) |  | 5 (10.2) |
| Native Hawaiian/Pacific Islander | 1 (0.5) | 2 (2.9) |  | 0 (0.0) |
| American Indian/Alaska Native | 4 (1.9) | 1 (1.4) |  | 0 (0.0) |
| Multi-racial | 5 (2.4) | 3 (4.3) |  | 1 (2.0) |
| Unknown | 25 (12.1) | 11 (15.7) |  | 5 (10.2) |
| Hispanic/Latino ethnicity, n (%) | 41 (19.8) | 18 (25.7) | 0.38 | 3 (6.1) |
| Comorbidities, n (%) | 86 (41.5) | 37 (52.9) | 0.13 | 22 (44.9) |
| Admission category, n (%) |  |  | <0.001* |  |
| Medical | 206 (99.5) | 52 (74.3) |  | 29 (59.2) |
| Surgical | 1 (0.5) | 10 (14.3) |  | 10 (20.4) |
| Trauma | 0 (0.0) | 8 (11.4) |  | 10 (20.4) |
| Antibiotics prior to intubation, n (%) | 72 (34.8) | 17 (24.3) | 0.14 | 17 (34.7) |
| Any antibiotic use, n (%) | 205 (99.0) | 64 (91.4) | 0.004* | 41 (83.7) |
| Ventilator days, median (IQR) | 7.0 (5.0-9.0) | 6.0 (4.0-7.0) | 0.003* | 6.0 (5.0-9.0) |
| ICU length of stay, median (IQR) | 11.0 (8.0-16.0) | 9.0 (7.0-12.0) | 0.009* | 10.0 (7.0-15.0) |
| Hospital length of stay, median (IQR) | 16.0 (11.0-24.0) | 17.0 (9.0-30.0) | 0.73 | 23.0 (14.0-43.0) |
| Mortality, n (%) | 6 (2.9) | 10 (14.3) | 0.001* | 2 (4.1) |

## SUPPLEMENTARY MATERIAL

### SUPPLEMENTARY FIGURES

**Figure S1.**
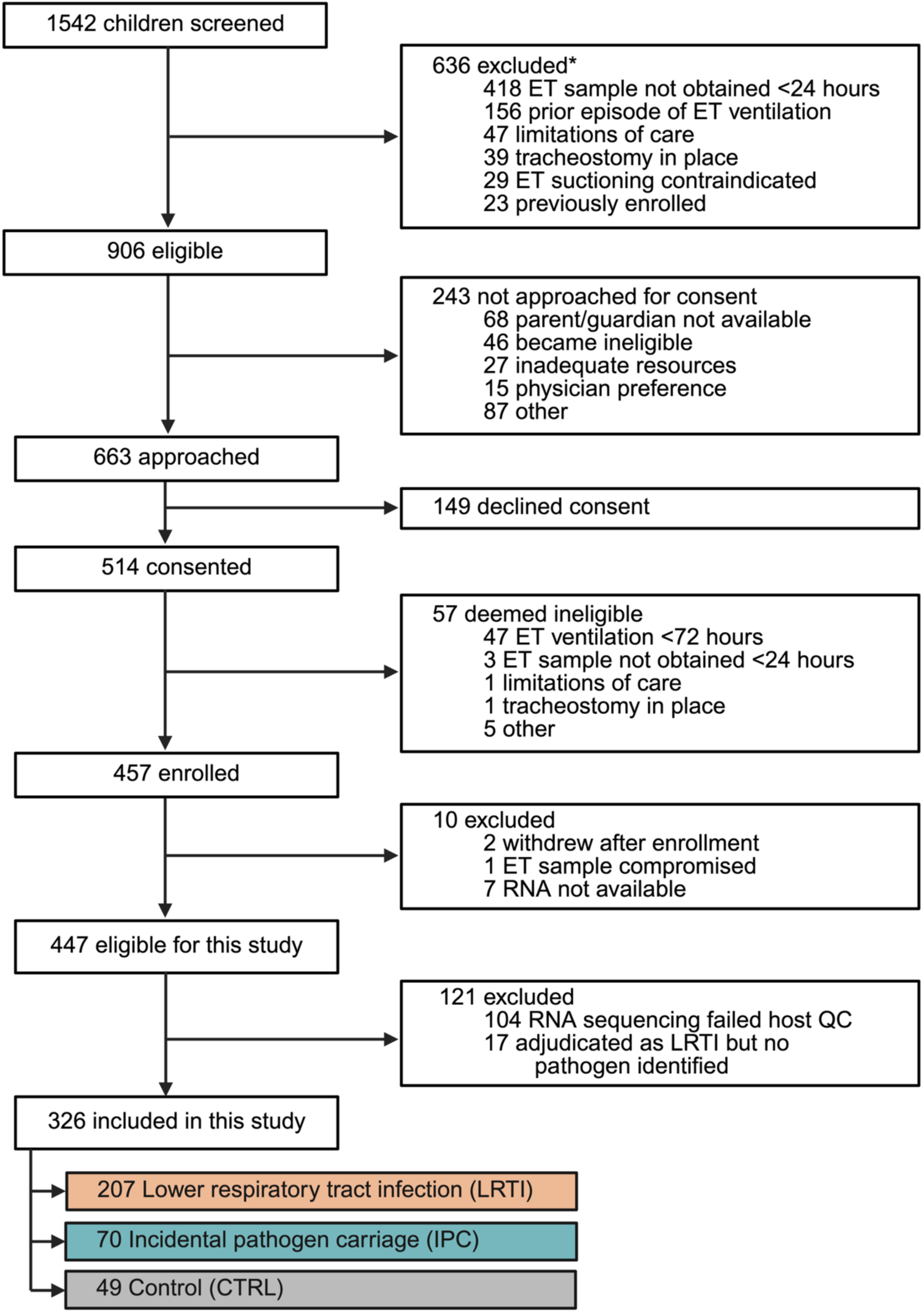
Study flow diagram. Asterisk indicates that some children had multiple reasons for exclusion. ET, endotracheal tube; QC, quality control.

**Figure S2.**
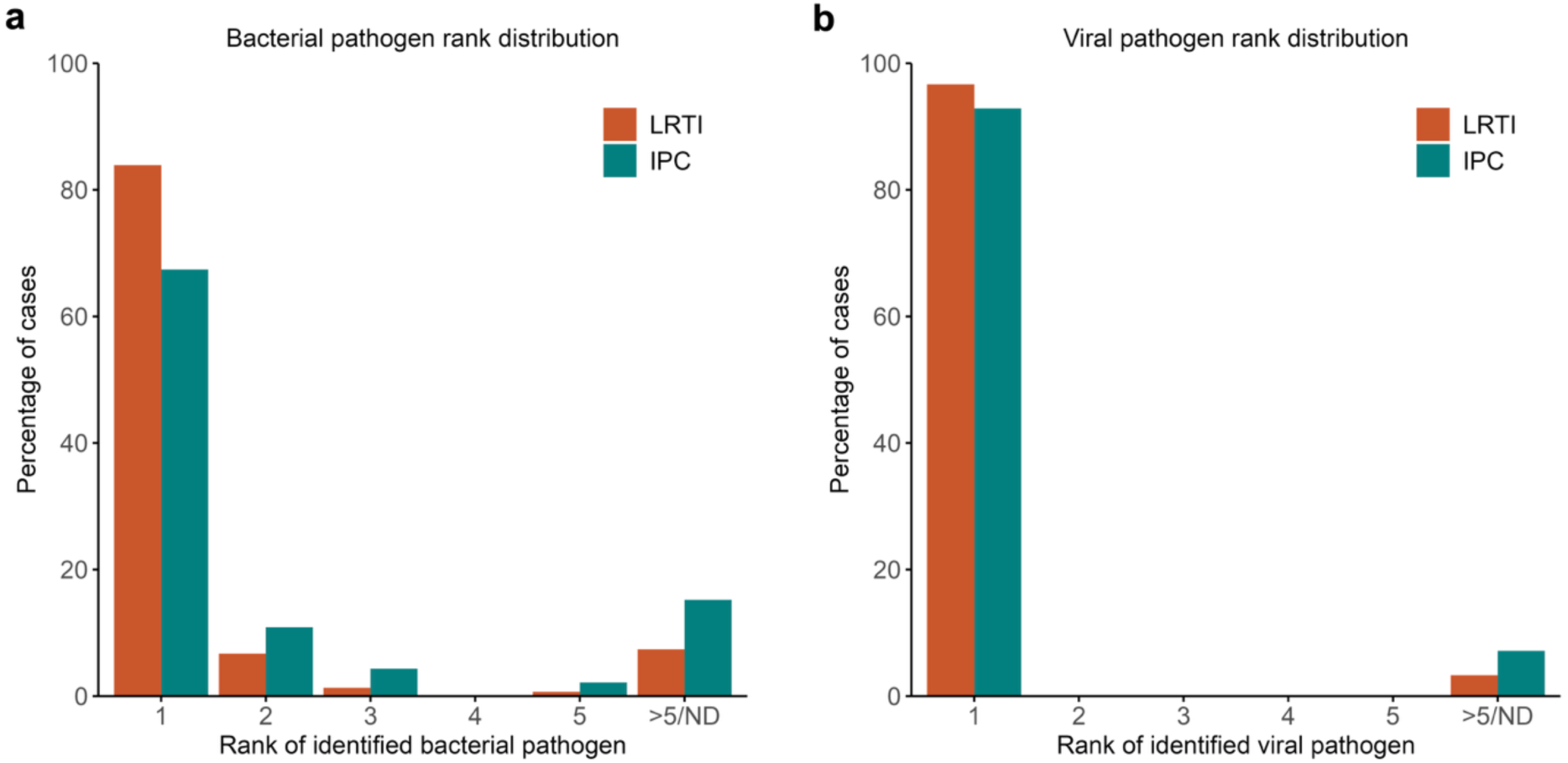
Ranks plots of detected bacterial and viral pathogens. **a.** Bar plot showing the distribution of pathogen ranks in patients who had a bacterial pathogen identified, stratified by LRTI and IPC groups. Rank was determined based on reads per million (RPM) of the detected pathogen in the metatranscriptomics data. If more than one bacterial pathogen was detected in a sample, the rank of the higher ranking pathogen was plotted. Ranks >5 and cases where the implicated pathogen was not identified in the metatranscriptomics data were grouped together as >5/ND. **b.** Bar plot showing the distribution of pathogen ranks in patients who had a viral pathogen identified, using the same methodology.

**Figure S3.**
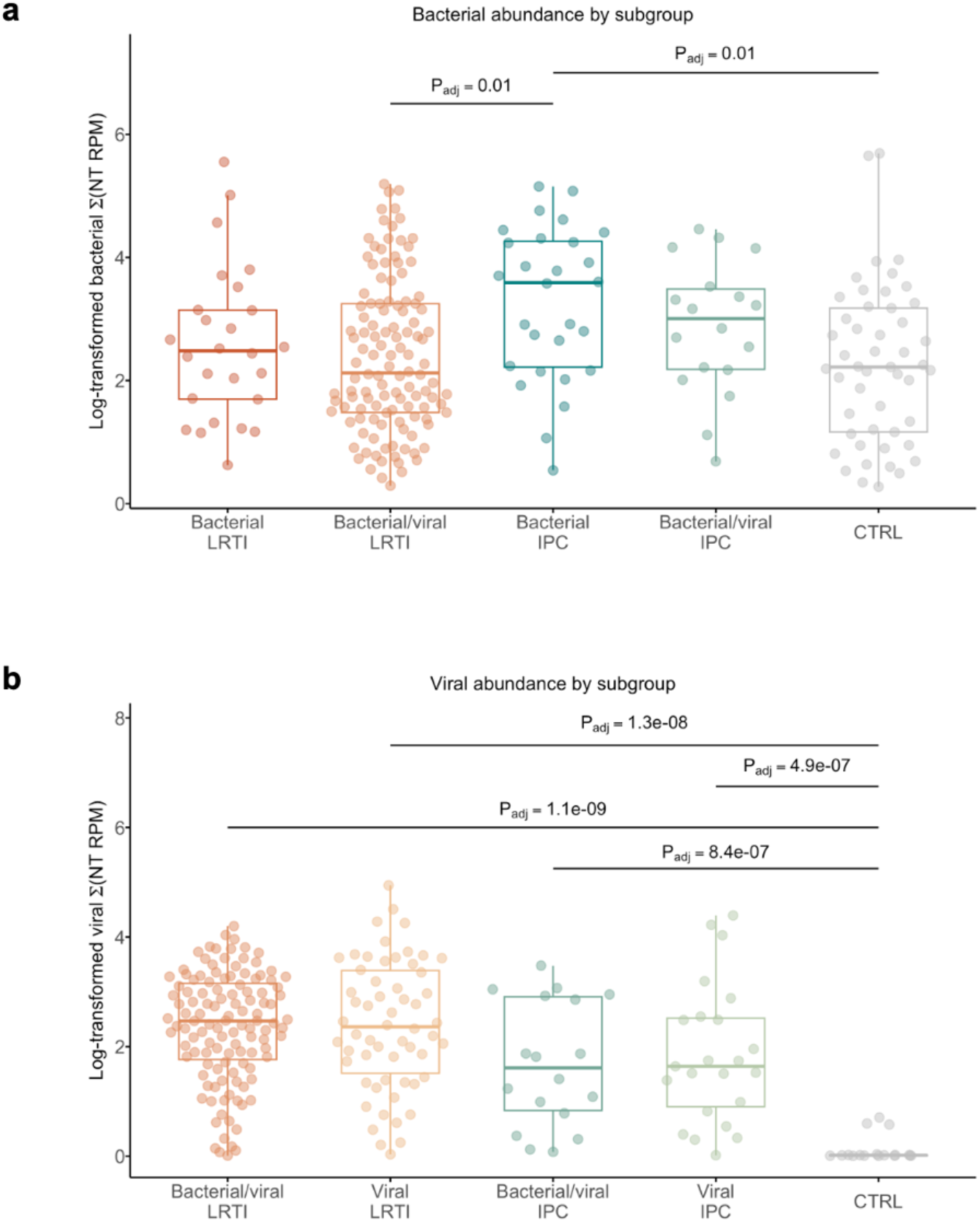
Microbial load by infection subclass. **a.** Total bacterial abundance measured in reads per million (RPM) across bacterial subclasses and controls. **b.** Total viral abundance across viral subclasses and controls. P values determined by Wilcoxon rank-sum tests and adjusted for multiple comparisons.

**Figure S4.**
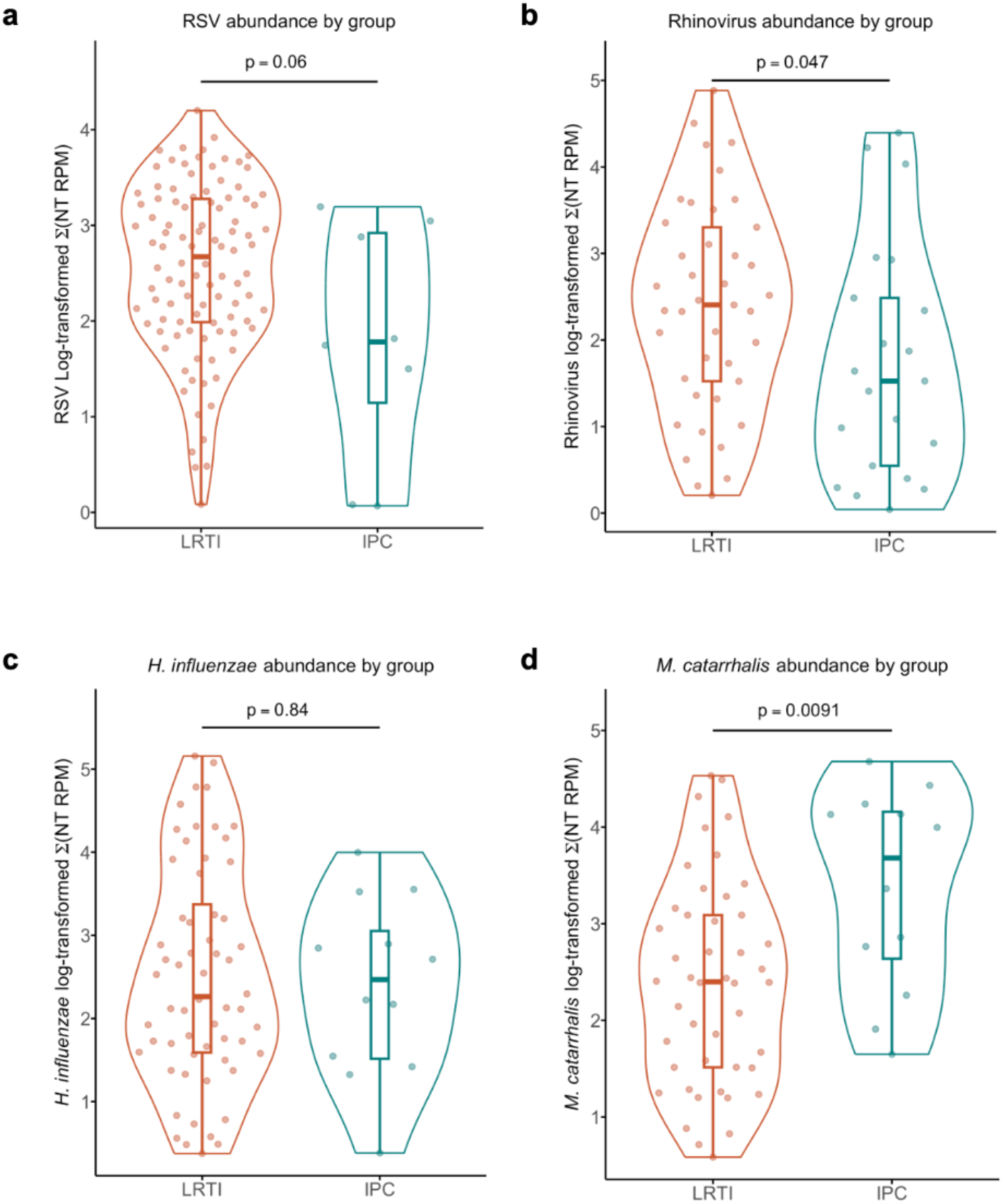
Abundance differences of the most prevalent bacterial and viral pathogens in LRTI versus IPC groups. **a.** Abundance of RSV, defined as sum of NT RPM, for patients who had RSV detected as a pathogen. **b.** Rhinovirus. **c.** *Haemophilus influenzae* **d.** *Moraxella catarrhalis*. NT RPM, nucleotide reads per million.

**Figure S5.**
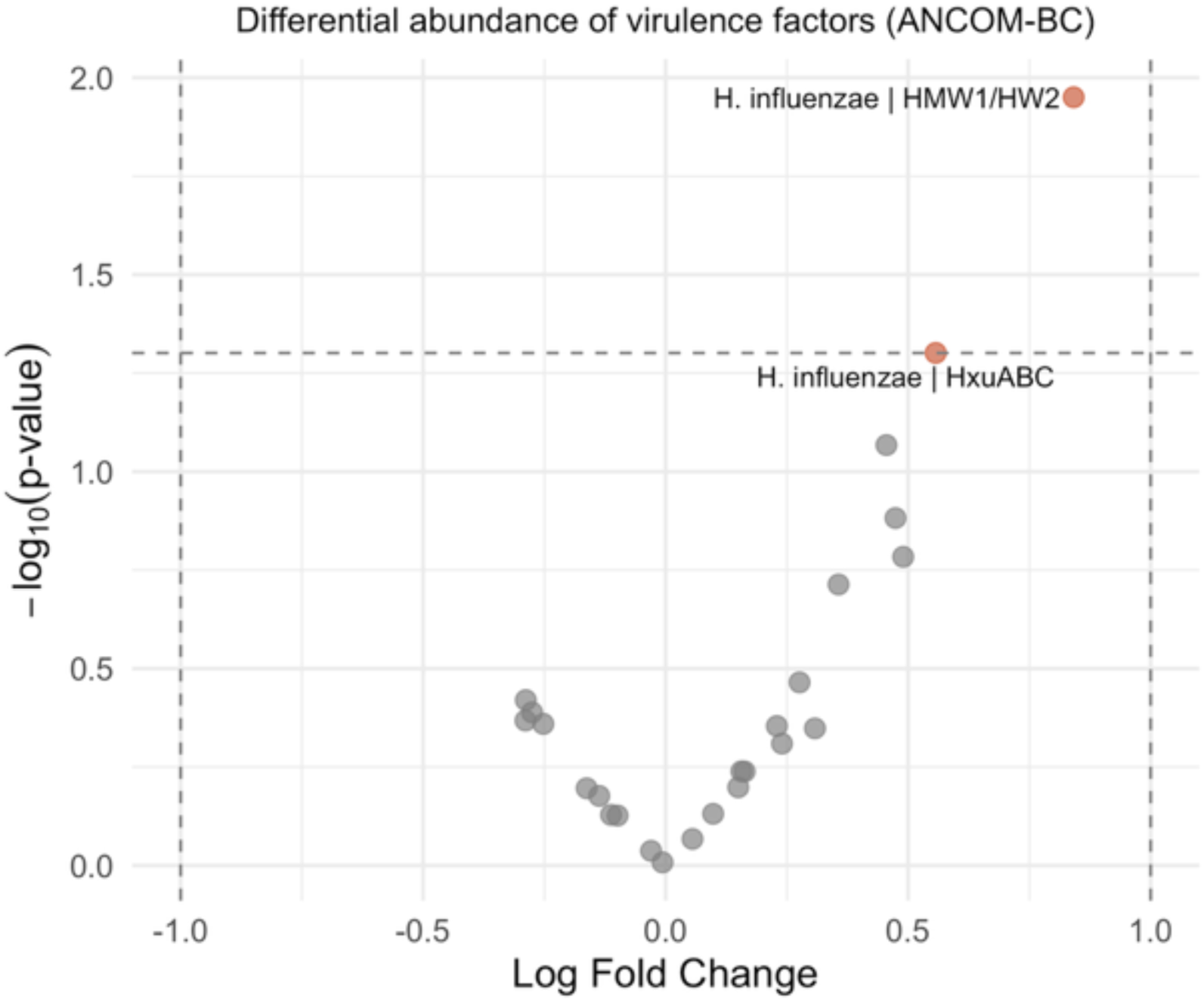
Exploratory analysis of virulence factors in LRTI and IPC groups. Differential abundance of all virulence factors, annotated by VFDB 2.0, between LRTI and IPC groups, using ANCOM-BC. Positive log fold change indicates upregulation with LRTI and negative log fold change indicates upregulation with IPC. Unadjusted P values shown.

**Figure S6.**
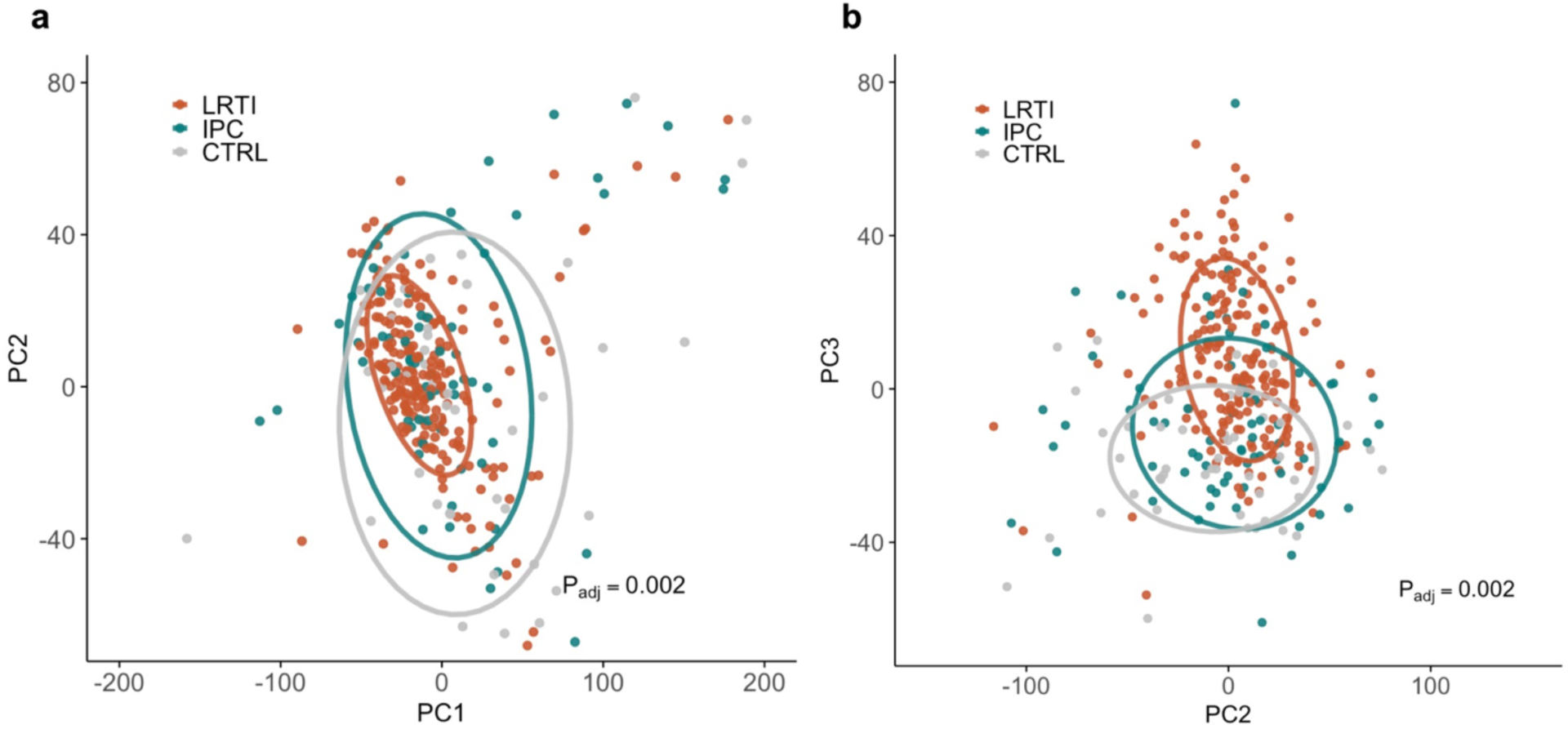
Additional principal components of the lower respiratory tract transcriptome. a. PC1 versus PC2. b. PC2 versus PC3. Orange indicates LRTI, teal indicates IPC, and gray indicates CTRL. Adjusted P values calculated by PERMANOVA between LRTI and non-LRTI groups.

**Figure S7.**
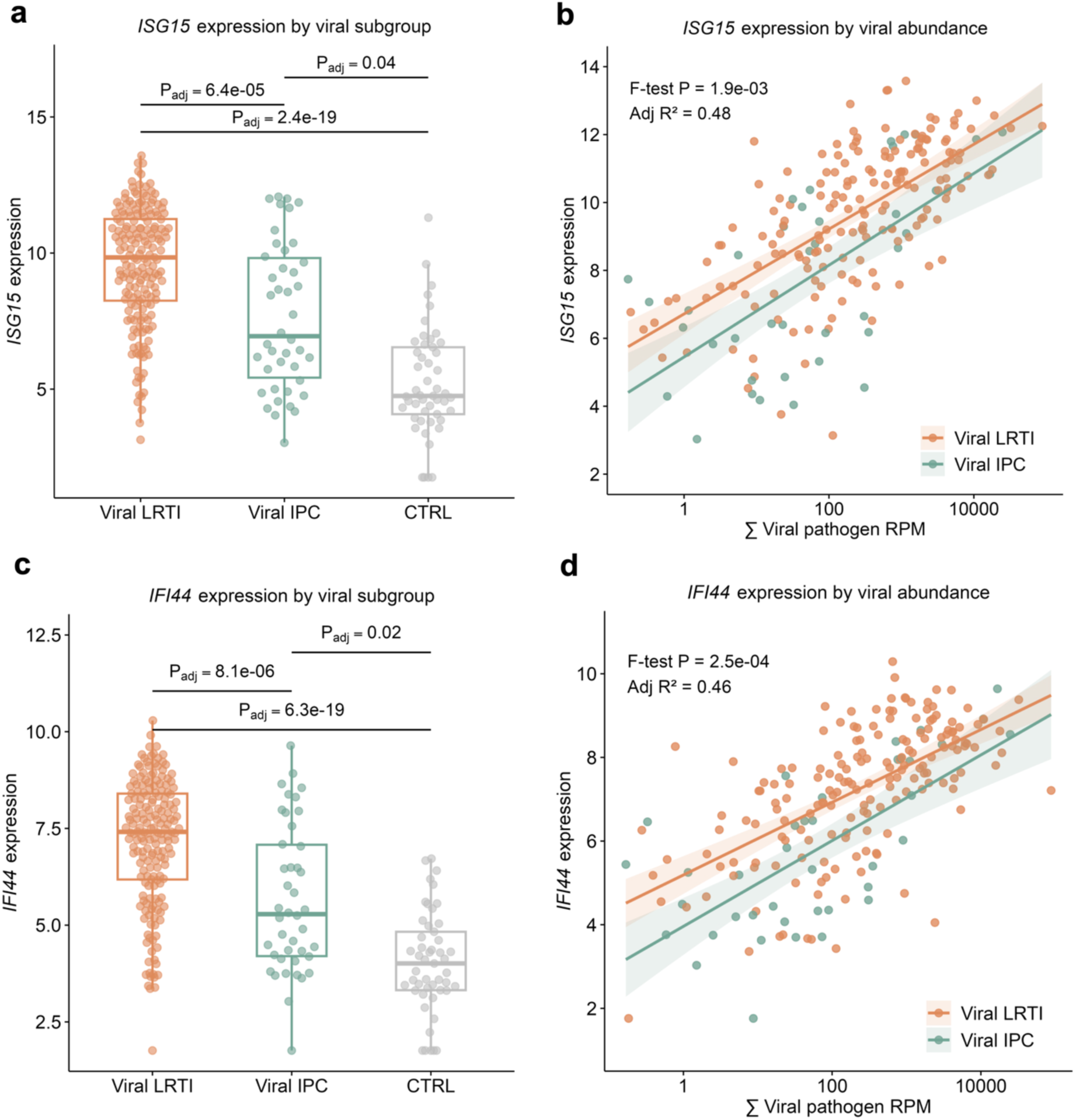
Integrated host-microbial analyses for additional interferon-related genes among viral cases. **a.** Boxplot of normalized *ISG15* expression by group. Benjamini-Hochberg (BH) adjusted P values, based on differential expression (DE) analyses, shown. **b.** Linear regression of viral abundance (reads per million, RPM), against normalized *ISG15* expression for viral LRTI and IPC. **c.** Boxplot of normalized *IFI44* expression by group. **d.** Linear regression of viral abundance against normalized *IFI44* expression for viral LRTI and IPC.

**Figure S8.**
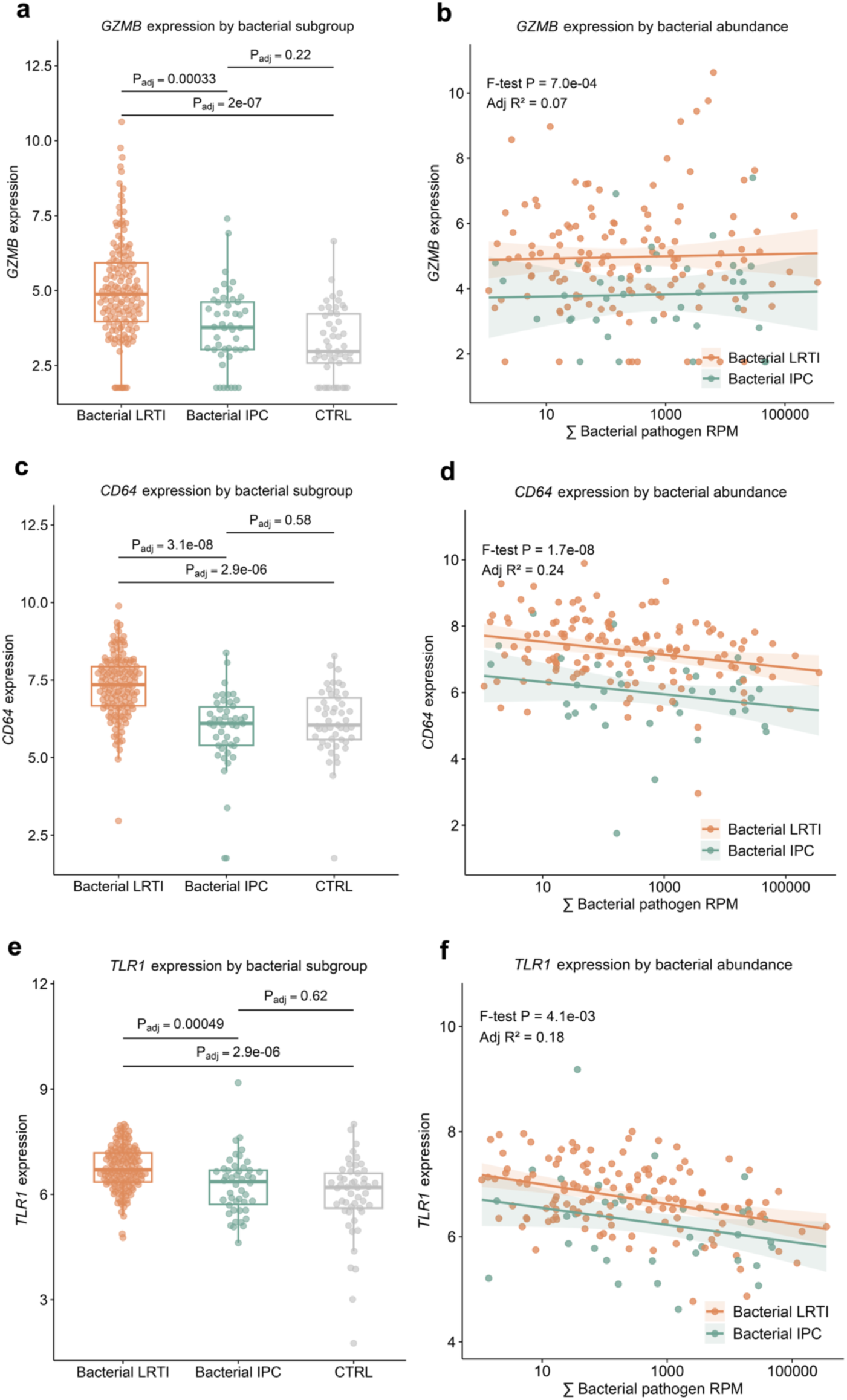
Integrated host-microbial analyses for classic anti-bacterial defense genes among bacterial cases. **a.** Boxplot of normalized *GZMB* expression. Benjamini-Hochberg (BH) adjusted P values, based on differential expression (DE) analyses, shown. **b.** Linear regression of bacterial pathogen abundance (reads per million, RPM), against normalized *GZMB* expression for bacterial LRTI and IPC. **c.** Boxplot of normalized *CD64* expression. **d.** Linear regression of bacterial abundance against normalized *CD64* expression for bacterial LRTI and IPC. **e.** Boxplot of normalized *TLR1* expression by group. **f.** Linear regression of bacterial abundance against normalized *TLR1* expression for bacterial LRTI and IPC.

### SUPPLEMENTARY TABLE

**Table S1.**
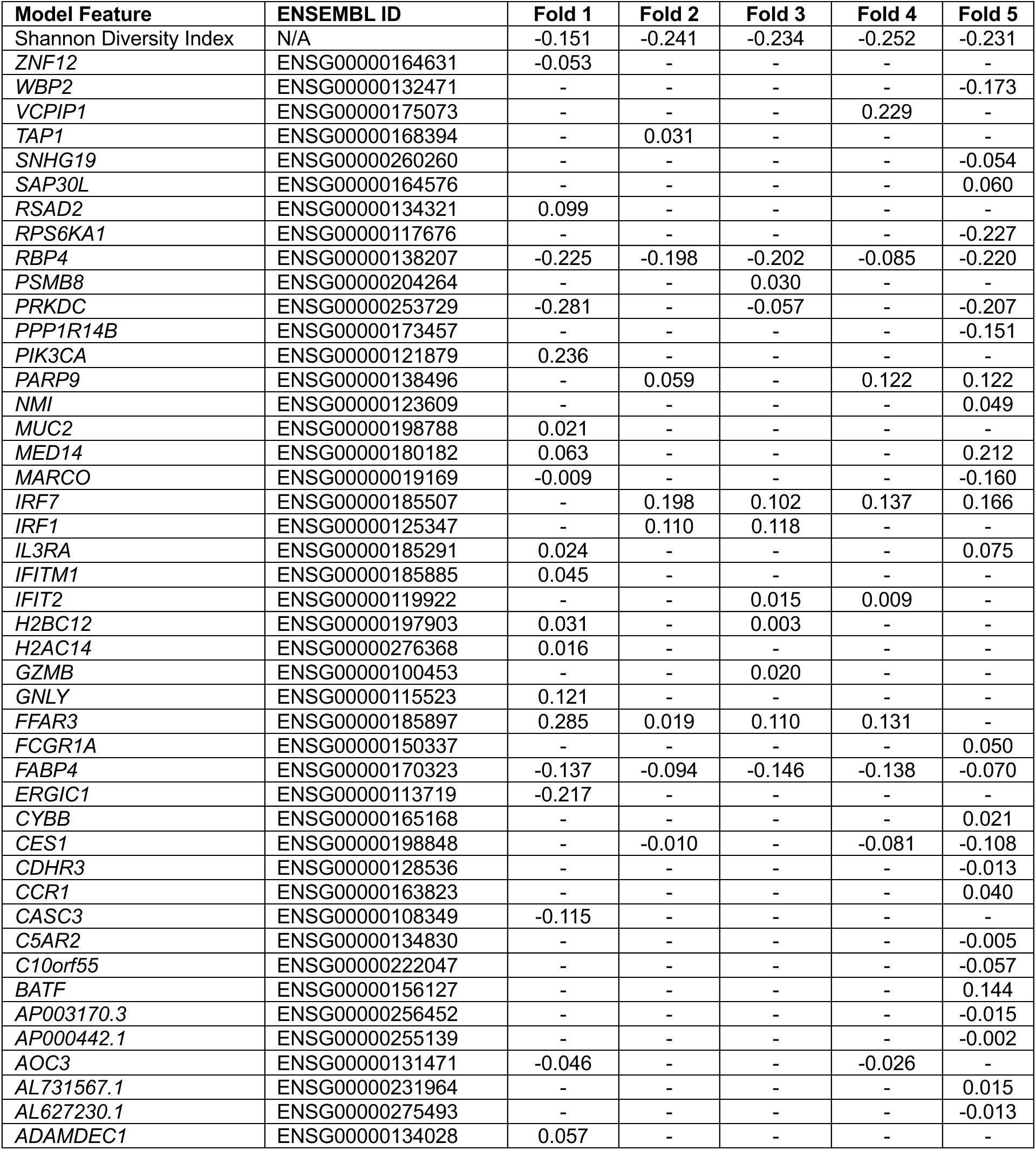
Integrated multi-gene and Shannon Diversity Index classifier coefficients. Model coefficients for each of the features included in the five folds are shown. If a gene was not included in that fold, its absence is indicated by a dash. Shannon diversity index, *FABP4*, and *RBP4*, were selected as model parameters in all folds.

### SUPPLEMENTARY DATA FILES

**Supplementary Data File 1:** ANCOM-BC of differential taxonomic abundance between LRTI and IPC, adjusted for age and sex. Legend: lfc = log fold change; se = standard error of log fold change; W = W test statistic; p = unadjusted P values; q = Benjamini-Hochberg adjusted P values; diff= denotes whether or not a given taxon is differentially abundant (TRUE/FALSE); passed_ss = denotes whether or not a given taxon passed structural zero screening (TRUE/FALSE).

**Supplementary Data File 2:** ANCOM-BC of differential virulence factor abundance between LRTI and IPC, adjusted for age and sex. Legend: lfc = log fold change; se = standard error of log fold change; W = W test statistic; p = unadjusted P values; q = Benjamini-Hochberg adjusted P values; diff= denotes whether or not a given taxon is differentially abundant (TRUE/FALSE); passed_ss = denotes whether or not a given taxon passed structural zero screening (TRUE/FALSE)

**Supplementary Data File 3:** Metabolic pathway differences between LRTI and IPC generated using HUMAnN3 and MaAsLin2. Legend: coef = effect size; stderr = standard error; N = number of samples; N.not.0 = number of samples with a value > 0; pval = unadjusted P values; qval = Benjamini-Hochberg adjusted P values.

**Supplementary Data File 4: a.** Genes differentially (DE) expressed between LRTI and CTRL, adjusted for age and sex. **b.** Genes DE between LRTI and IPC, adjusted for age and sex. **c.** Genes DE between IPC and CTRL, adjusted for age and sex. Legend: logFC = log_2_ fold change; P_adj_ = Benjamini-Hochberg adjusted P values, t = T statistic, B = B statistic.

**Supplementary Data File 5: a.** Gene set enrichment analyses (GSEA) of DE genes between LRTI and IPC. **b.** GSEA of DE genes between IPC and CTRL. Legend: NES = normalized enrichment score, p.adjust = Benjamini-Hochberg adjusted P value, setSize = size of REACTOME pathway.

**Supplementary Data File 6: a.** Genes DE between viral LRTI and CTRL, adjusted for age and sex. **b.** Genes DE between bacterial LRTI and CTRL, adjusted for age and sex. **c.** Genes DE between viral LRTI and viral IPC, adjusted for age and sex. **d.** Genes DE between bacterial LRTI and bacterial IPC, adjusted for age and sex. **e.** Genes DE between viral IPC and CTRL, adjusted for age and sex. **f.** Genes DE between bacterial IPC and CTRL, adjusted for age and sex. Legend: logFC = log_2_ fold change; P_adj_ = Benjamini-Hochberg adjusted P values, t = T statistic, B = B statistic.

**Supplementary Data File 7: a.** Genes differentially (DE) expressed between LRTI and CTRL, adjusted for age, sex, and Shannon Diversity Index (SDI). **b.** Genes DE between LRTI and IPC, adjusted for age, sex, and SDI. **c.** Genes DE between IPC and CTRL, adjusted for age, sex, and SDI. Legend: logFC = log_2_ fold change; P_adj_ = Benjamini-Hochberg adjusted P values, t = T statistic, B = B statistic.

## REFERENCES

1. Man, W. H., de Steenhuijsen Piters, W. A. A. & Bogaert, D. The microbiota of the respiratory tract: gatekeeper to respiratory health. Nat Rev Microbiol 15, 259–270 (2017).

2. Robinson, J. Colonization and infection of the respiratory tract: What do we know? Paediatr Child Health 9, 21–24 (2004).

3. Vo, P. & Kharasch, V. S. Respiratory Failure. Pediatrics In Review 35, 476–486 (2014).

4. Panetti, B. et al. Acute Respiratory Failure in Children: A Clinical Update on Diagnosis. Children 11, 1232 (2024).

5. Jeffrey, M., Denny, K. J., Lipman, J. & Conway Morris, A. Differentiating infection, colonisation, and sterile inflammation in critical illness: the emerging role of host-response profiling. Intensive Care Med 49, 760–771 (2023).

6. Lydon, E. C., Ko, E. R. & Tsalik, E. L. The host response as a tool for infectious disease diagnosis and management. Expert Review of Molecular Diagnostics 18, 723–738 (2018).

7. Vaughn, V. M. et al. Excess Antibiotic Treatment Duration and Adverse Events in Patients Hospitalized With Pneumonia: A Multihospital Cohort Study. Ann Intern Med 171, 153–163 (2019).

8. Gupta, A. B. et al. Inappropriate Diagnosis of Pneumonia Among Hospitalized Adults. JAMA Intern Med 184, 548–556 (2024).

9. Anadol, D., Aydin, Y. Z. & Göçmen, A. Overdiagnosis of pneumonia in children. Turk J Pediatr 43, 205–209 (2001).

10. Pan, H., Cui, B., Huang, Y., Yang, J. & Ba-Thein, W. Nasal carriage of common bacterial pathogens among healthy kindergarten children in Chaoshan region, southern China: a cross-sectional study. BMC Pediatr 16, 161 (2016).

11. Regev-Yochay, G. et al. Nasopharyngeal Carriage of *Streptococcus pneumoniae* by Adults and Children in Community and Family Settings. CLIN INFECT DIS 38, 632–639 (2004).

12. Nokso-Koivisto, J., Kinnari, T. J., Lindahl, P., Hovi, T. & Pitkäranta, A. Human picornavirus and coronavirus RNA in nasopharynx of children without concurrent respiratory symptoms. J Med Virol 66, 417–420 (2002).

13. Parker, A. M. et al. Upper respiratory *Streptococcus pneumoniae* colonization among working-age adults with prevalent exposure to overcrowding. Microbiol Spectr 12, e00879–24 (2024).

14. Desai, A. P. et al. Decline in Pneumococcal Nasopharyngeal Carriage of Vaccine Serotypes After the Introduction of the 13-Valent Pneumococcal Conjugate Vaccine in Children in Atlanta, Georgia. Pediatr Infect Dis J 34, 1168–1174 (2015).

15. Bogaert, D., De Groot, R. & Hermans, P. W. M. Streptococcus pneumoniae colonisation: the key to pneumococcal disease. Lancet Infect Dis 4, 144–154 (2004).

16. Huang, S. S. et al. Healthcare utilization and cost of pneumococcal disease in the United States. Vaccine 29, 3398–3412 (2011).

17. Vaneechoutte, M., Verschraegen, G., Claeys, G., Weise, B. & Van den Abeele, A. M. Respiratory tract carrier rates of Moraxella (Branhamella) catarrhalis in adults and children and interpretation of the isolation of M. catarrhalis from sputum. J Clin Microbiol 28, 2674– 2680 (1990).

18. Faden, H., Harabuchi, Y. & Hong, J. J. Epidemiology of Moraxella catarrhalis in children during the first 2 years of life: relationship to otitis media. J Infect Dis 169, 1312–1317 (1994).

19. Ejlertsen, T., Thisted, E., Ebbesen, F., Olesen, B. & Renneberg, J. Branhamella catarrhalis in children and adults. A study of prevalence, time of colonisation, and association with upper and lower respiratory tract infections. J Infect 29, 23–31 (1994).

20. Most, Z. M., Perl, T. M. & Sebert, M. Respiratory virus infections in symptomatic and asymptomatic children upon hospital admission: new insights. Antimicrob Steward Healthc Epidemiol 4, e162 (2024).

21. Jansen, R. R. et al. Frequent Detection of Respiratory Viruses without Symptoms: Toward Defining Clinically Relevant Cutoff Values ▿. J Clin Microbiol 49, 2631–2636 (2011).

22. Self, W. H. et al. Respiratory Viral Detection in Children and Adults: Comparing Asymptomatic Controls and Patients With Community-Acquired Pneumonia. J Infect Dis 213, 584–591 (2016).

23. Dickson, R. P. The microbiome and critical illness. The Lancet Respiratory Medicine 4, 59– 72 (2016).

24. Mourani, P. M. et al. Temporal airway microbiome changes related to ventilator-associated pneumonia in children. European Respiratory Journal 57, (2021).

25. Durairaj, L. et al. Patterns and density of early tracheal colonization in intensive care unit patients. J Crit Care 24, 114–121 (2009).

26. Ewig, S. et al. Bacterial colonization patterns in mechanically ventilated patients with traumatic and medical head injury. Incidence, risk factors, and association with ventilator-associated pneumonia. Am J Respir Crit Care Med 159, 188–198 (1999).

27. Mick, E. et al. Integrated host/microbe metagenomics enables accurate lower respiratory tract infection diagnosis in critically ill children. J Clin Invest 133, (2023).

28. Tsitsiklis, A. et al. Lower respiratory tract infections in children requiring mechanical ventilation: a multicentre prospective surveillance study incorporating airway metagenomics. The Lancet Microbe 3, e284–e293 (2022).

29. Lydon, E., et al. Proteomic profiling of the local and systemic immune response to pediatric respiratory viral infections. mSystems 10, e0133524 (2025).

30. United States Centers for Disease Control and Prevention. CDC/NHSN Surveillance Definitions for Specific Types of Infections. https://www.cdc.gov/nhsn/pdfs/pscmanual/pcsmanual_current.pdf (2021).

31. Patel, R. et al. Clinically Adjudicated Reference Standards for Evaluation of Infectious Diseases Diagnostics. Clin Infect Dis 76, 938–943 (2023).

32. Nitu, M. E. & Eigen, H. Respiratory Failure. Pediatrics In Review 30 470–478 (2009).

33. Lin, H. & Peddada, S. D. Analysis of compositions of microbiomes with bias correction. Nat Commun 11, 3514 (2020).

34. Dong, W. et al. An expanded database and analytical toolkit for identifying bacterial virulence factors and their associations with chronic diseases. Nat Commun 15, 8084 (2024).

35. St Geme, J. W. & Yeo, H.-J. A prototype two-partner secretion pathway: the Haemophilus influenzae HMW1 and HMW2 adhesin systems. Trends Microbiol 17, 355–360 (2009).

36. Akhtar, A. A. & Turner, D. PJ. The role of bacterial ATP-binding cassette (ABC) transporters in pathogenesis and virulence: Therapeutic and vaccine potential. Microbial Pathogenesis 171, 105734 (2022).

37. Beghini, F. et al. Integrating taxonomic, functional, and strain-level profiling of diverse microbial communities with bioBakery 3. eLife 10, e65088 (2021).

38. Furuhashi, M., Saitoh, S., Shimamoto, K. & Miura, T. Fatty Acid-Binding Protein 4 (FABP4): Pathophysiological Insights and Potent Clinical Biomarker of Metabolic and Cardiovascular Diseases. Clin Med Insights Cardiol 8, 23–33 (2014).

39. Schneider, W. M., Chevillotte, M. D. & Rice, C. M. Interferon-Stimulated Genes: A Complex Web of Host Defenses. Annu Rev Immunol 32, 513–545 (2014).

40. Hofer, U. Granzyme B’s roundhouse kick against bacteria. Nat Rev Microbiol 15, 707–707 (2017).

41. Icardi, M. et al. CD64 Index Provides Simple and Predictive Testing for Detection and Monitoring of Sepsis and Bacterial Infection in Hospital Patients. J Clin Microbiol 47, 3914– 3919 (2009).

42. Albiger, B., Dahlberg, S., Henriques-Normark, B. & Normark, S. Role of the innate immune system in host defence against bacterial infections: focus on the Toll-like receptors. J Intern Med 261, 511–528 (2007).

43. Lydon, E. C. et al. Pulmonary *FABP4* Is an Inverse Biomarker of Pneumonia in Critically Ill Children and Adults. Am J Respir Crit Care Med 210, 1480–1483 (2024).

44. Langelier, C. et al. Integrating host response and unbiased microbe detection for lower respiratory tract infection diagnosis in critically ill adults. Proc Natl Acad Sci U S A 115, E12353–E12362 (2018).

45. Flanagan, J. L. et al. Loss of bacterial diversity during antibiotic treatment of intubated patients colonized with Pseudomonas aeruginosa. J. Clin. Microbiol. 45, 1954–1962 (2007).

46. Wesolowska-Andersen, A. et al. Dual RNA-seq reveals viral infections in asthmatic children without respiratory illness which are associated with changes in the airway transcriptome. Genome Biol 18, 12 (2017).

47. Wolsk, H. M. et al. Picornavirus-Induced Airway Mucosa Immune Profile in Asymptomatic Neonates. J Infect Dis 213, 1262–1270 (2016).

48. Følsgaard, N. V. et al. Pathogenic bacteria colonizing the airways in asymptomatic neonates stimulates topical inflammatory mediator release. Am J Respir Crit Care Med 187, 589–595 (2013).

49. Bisgaard, H. et al. Childhood asthma after bacterial colonization of the airway in neonates. N Engl J Med 357, 1487–1495 (2007).

50. Cho, H.-J. et al. Differences and similarities between the upper and lower airway: focusing on innate immunity. Rhinology 59, 441–450 (2021).

51. Mick, E. et al. Upper airway gene expression reveals suppressed immune responses to SARS-CoV-2 compared with other respiratory viruses. Nat Commun 11, 5854 (2020).

52. Mick, E. et al. Upper airway gene expression shows a more robust adaptive immune response to SARS-CoV-2 in children. Nat Commun 13, 3937 (2022).

53. Ichinohe, T. et al. Microbiota regulates immune defense against respiratory tract influenza A virus infection. Proc. Natl. Acad. Sci. U.S.A. 108, 5354–5359 (2011).

54. Vanderweele, T. J. & Vansteelandt, S. Conceptual issues concerning mediation, interventions and composition. Statistics and Its Interface 2, 457–468 (2009).

55. Gaston, D. C. Clinical Metagenomics for Infectious Diseases: Progress toward Operational Value. J Clin Microbiol 61, e01267–22 (2023).

56. Benoit, P. et al. Seven-year performance of a clinical metagenomic next-generation sequencing test for diagnosis of central nervous system infections. Nat Med 30, 3522–3533 (2024).

57. National Human Genome Research Institute. DNA Sequencing Costs: Data. https://www.genome.gov/about-genomics/fact-sheets/DNA-Sequencing-Costs-Data (2024).

58. Self, W. H. et al. Procalcitonin as a Marker of Etiology in Adults Hospitalized With Community-Acquired Pneumonia. Clin Infect Dis 65, 183–190 (2017).

59. van der Meer, V., Neven, A. K., van den Broek, P. J. & Assendelft, W. J. J. Diagnostic value of C reactive protein in infections of the lower respiratory tract: systematic review. BMJ 331, 26 (2005).

60. Gu, W. et al. Depletion of Abundant Sequences by Hybridization (DASH): using Cas9 to remove unwanted high-abundance species in sequencing libraries and molecular counting applications. Genome Biology 17, 1–13 (2016).

61. Gold, L. et al. Aptamer-based multiplexed proteomic technology for biomarker discovery. PLoS One 5, e15004 (2010).

62. Kim, C. H. et al. Stability and reproducibility of proteomic profiles measured with an aptamer-based platform. Sci Rep 8, 8382 (2018).

63. Candia, J. et al. Variability of 7K and 11K SomaScan Plasma Proteomics Assays. J Proteome Res 23, 5531–5539 (2024).

64. Kalantar, K. L. et al. IDseq-An open source cloud-based pipeline and analysis service for metagenomic pathogen detection and monitoring. Gigascience 9, giaa111 (2020).

65. Lu, D. et al. Simultaneous detection of pathogens and antimicrobial resistance genes with the open source, cloud-based, CZ ID platform. Genome Med 17, 46 (2025).

66. Bray, N. L., Pimentel, H., Melsted, P. & Pachter, L. Near-optimal probabilistic RNA-seq quantification. Nature Biotechnology 34, 525–527 (2016).

67. Chen, S., Zhou, Y., Chen, Y. & Gu, J. fastp: an ultra-fast all-in-one FASTQ preprocessor. Bioinformatics 34, i884–i890 (2018).

68. Ruby, J. G., Bellare, P. & Derisi, J. L. PRICE: software for the targeted assembly of components of (Meta) genomic sequence data. G3 (Bethesda) 3, 865–880 (2013).

69. Langmead, B. & Salzberg, S. L. Fast gapped-read alignment with Bowtie 2. Nat Methods. 9, (2012).

70. Jain, S. et al. Community-Acquired Pneumonia Requiring Hospitalization among U.S. Adults. New England Journal of Medicine 373, 415–427 (2015).

71. Jain, S. et al. Community-acquired pneumonia requiring hospitalization among U.S. children. N Engl J Med 372, 835–845 (2015).

72. Iwai, S. et al. The Lung Microbiome of Ugandan HIV-Infected Pneumonia Patients Is Compositionally and Functionally Distinct from That of San Franciscan Patients. PLOS ONE 9, e95726 (2014).

73. Magill, S. S. et al. Multistate point-prevalence survey of health care-associated infections. N Engl J Med 370, 1198–1208 (2014).

74. Soneson, C., Love, M. I. & Robinson, M. D. Differential analyses for RNA-seq: transcript-level estimates improve gene-level inferences. F1000Res 4, 1521 (2015).

75. Love, M. I., Huber, W. & Anders, S. Moderated estimation of fold change and dispersion for RNA-seq data with DESeq2. Genome Biol 15, 550 (2014).

76. Ritchie, M. E. et al. limma powers differential expression analyses for RNA-sequencing and microarray studies. Nucleic Acids Res 43, e47 (2015).

77. Croft, D. et al. The Reactome pathway knowledgebase. Nucleic Acids Res 42, D472–D477 (2014).

78. Tingley, D., Yamamoto, T., Hirose, K., Keele, L. & Imai, K. mediation: R Package for Causal Mediation Analysis. Journal of Statistical Software 59, 1–38 (2014).

79. Friedman, J., et al. glmnet: Lasso and Elastic-Net Regularized Generalized Linear Models. (2023).

80. Robin, X. et al. pROC: an open-source package for R and S+ to analyze and compare ROC curves. BMC Bioinformatics 12, 77 (2011).

